# GWAS of macro-scale resting state functional brain networks identify shared biology with brain structure and autism

**DOI:** 10.1101/2025.04.27.25326490

**Authors:** Yuankai He, Rafael Romero-Garcia, Bin Wan, Jakob Grove, Anders D. Børglum, Simon Baron-Cohen, Sofie L. Valk, Edward T. Bullmore, Richard A.I. Bethlehem, Varun Warrier

**Affiliations:** Department of Psychiatry, University of Cambridge, Sir William Hardy Building, Downing Site, CB2 3EB, Cambridge, UK; Instituto de Biomedicina de Sevilla (IBiS) HUVR/CSIC/Universidad de Sevilla/CIBERSAM, ISCIII, Dpto. de Fisiología Médica y Biofísica, Sevilla, Spain; Institute of Neuroscience and Medicine (INM-7: Brain and Behavior), Research Center Jülich, Jülich, Germany; Max-Planck-Institut für Kognitions- und Neurowissenschaften, Stephanstraße 1a, 04103 Leipzig, Germany; University Hospitals of Geneva, Geneva, Switzerland; The Lundbeck Foundation Initiative for Integrative Psychiatric Research, iPSYCH, Aarhus, 8010, Denmark; Center for Genomics and Personalized Medicine (CGPM), Aarhus University, Aarhus, 8000, Denmark; Department of Biomedicine (Human Genetics) and iSEQ Center, Aarhus University, Aarhus, 8000, Denmark; Bioinformatics Research Centre, Aarhus University, Aarhus, Denmark, 8000; Department of Psychology, University of Cambridge, Downing Street, CB2 3EB, Cambridge, UK

**Author notes:** Equal contribution, order can be reversed in citations (Richard A.I. Bethlehem, Varun Warrier).

## Abstract

The topology of the functional brain network has been associated with several neuropsychiatric conditions. However, little is known about its genetic underpinnings, and whether this overlaps with brain structure and neuropsychiatric conditions. Hence, we conducted genome-wide association study across six graph metrics of the macroscale functional networks at global, hemispheric and regional levels and their hemispheric asymmetry in 54,030 individuals. We identified seven experiment-wide significant loci and prioritised ten candidate genes. Both global graph metrics and hemispheric asymmetry are modestly heritable, but phenotypically and genetically form two clusters. Furthermore, cortical macro- and microstructure are causally related to global graph metrics and asymmetry, respectively, suggesting a dual structural constraint on functional network organisation. Finally, amongst twelve common neuropsychiatric conditions, only autism was genetically correlated with graph metrics of the functional network, supporting phenotypic case-control differences in functional connectivity. Overall, our results suggest different genetic axes shaping different aspects of brain functional topology and demonstrate shared biology with brain structure and autism.

## 1 Introduction

Spontaneous fluctuations of brain activity have been a topical area of research since it was observed that different brain regions display temporally coordinated activity even in the absence of extrinsic stimuli [1]. Subsequent analyses have identified reproducible patterns of closely correlated temporal dynamics within functionally related brain regions [2–4], whose topology can be modelled using a network of nodes (brain regions) and edges (functional connectivity between brain regions) [5, 6]. Using mathematical principles derived from graph theory, complex organisational properties of brain function can be quantified with graph metrics. Important features include organisation of brain regions into distinct functional communities (modularity), balance between segregation and integration (small-worldness), and a core set of densely connected hub regions (rich-club structure) [6–10]. Because resting-state functional magnetic resonance imaging (fMRI) can be used across different age groups [11, 12] and levels of cognitive functioning [13], decades of research have linked resting-state fMRI networks with cognitive function, mental health, and neuropathologies [14–17]. In particular, overarching topological properties of the functional network have been hypothesised to be a converging feature across different neuropsychiatric conditions. [18].

Despite phenotypic theories linking functional network organisation with various neuropsychiatric conditions, much less is known about the genetic ontology underlying functional network organisation, and if it is shared with neuropsychiatric conditions. Twin-based and initial genome-wide association study (GWAS) findings have shown that the resting-state connectome has modest but significant heritability [19–24], with some overlapping loci and genes with mental health conditions [25]. However, initial GWAS results for regional connectivity strengths [22], functional network level independent component-based signatures [21] and global efficiency [23] have been heterogenous, showing only two shared gene-level associations and no overlapping disease correlates. It is unclear to what extent the genetic underpinnings are shared between global and regional network architecture. Also, whilst valuable genetic knowledge has been gained from studies using functional networks averaged across hemispheres, it remains unknown if hemispheric asymmetry of functional networks is driven by separate biological processes from global functional topology. This can be clinically important as numerous neuropsychiatric conditions have been phenotypically associated with functional network asymmetry [26, 27], and genetically correlated with structural asymmetry [28]. Hence, the first major aim of this study was to systematically characterise the genetic processes underlying global, regional and asymmetric organisation of the functional network.

Another incompletely resolved question is the relationship between functional and structural organisation of the human brain. The conventional view suggests that functional interactions between different brain regions requires anatomical connections between the neuronal populations in these regions [29, 30]. Hence, functional network topology is suggested to be constrained by the white matter connectome, which can be measured by diffusion MRI [31]. A competing view instead understands brain activity fluctuations as standing waves, and posits that spatiotemporal coordination of brain activity is constrained by the macrostructural geometry of the brain [32]. Whilst both theories have some empirical support, it cannot be determined based on only neuroimaging evidence which structural phenotypes are biologically related or even causal to functional network architecture. The recent availability of genetic findings for brain structure [33] allowed us to pursue the second major aim: to identify genetic correlations between brain structure and function, and infer causality using Mendelian Randomisation [34].

Finally, whilst both structural and functional differences have been associated with neuropsychiatric conditions, individual-level predictors derived from resting-state connectomics only show partial overlap with those identified from structural neuroimaging [35, 36]. Hence, the third major aim of this study was to identify the overlapping genetic processes underlying functional network topology and neuropsychiatric conditions, and to test if these overlaps were independent from brain structure.

To achieve these three aims, we derived seven graph theory metrics [37] from resting-state fMRI at global, hemispheric and regional levels based on the most fine-grained multimodal parcellation atlas to date [38]. Because multiple measures of asymmetry have been studied phenotypically [39–42] but none in a GWAS study, we used three different measures of hemispheric asymmetry to select most heritable measure. For these 2,291 phenotypes (7 global, 10 hemispheric, 2,256 regional, 6 asymmetry × 3 parametrisations), we conducted GWAS in 54,030 individuals from the UK Biobank, estimated their heritability, explored gene- and pathway-level associations, and investigated their shared genetics and possible causal relationships with brain structure as well as neuropsychiatric conditions. (Figure 1)

**Figure 1:**
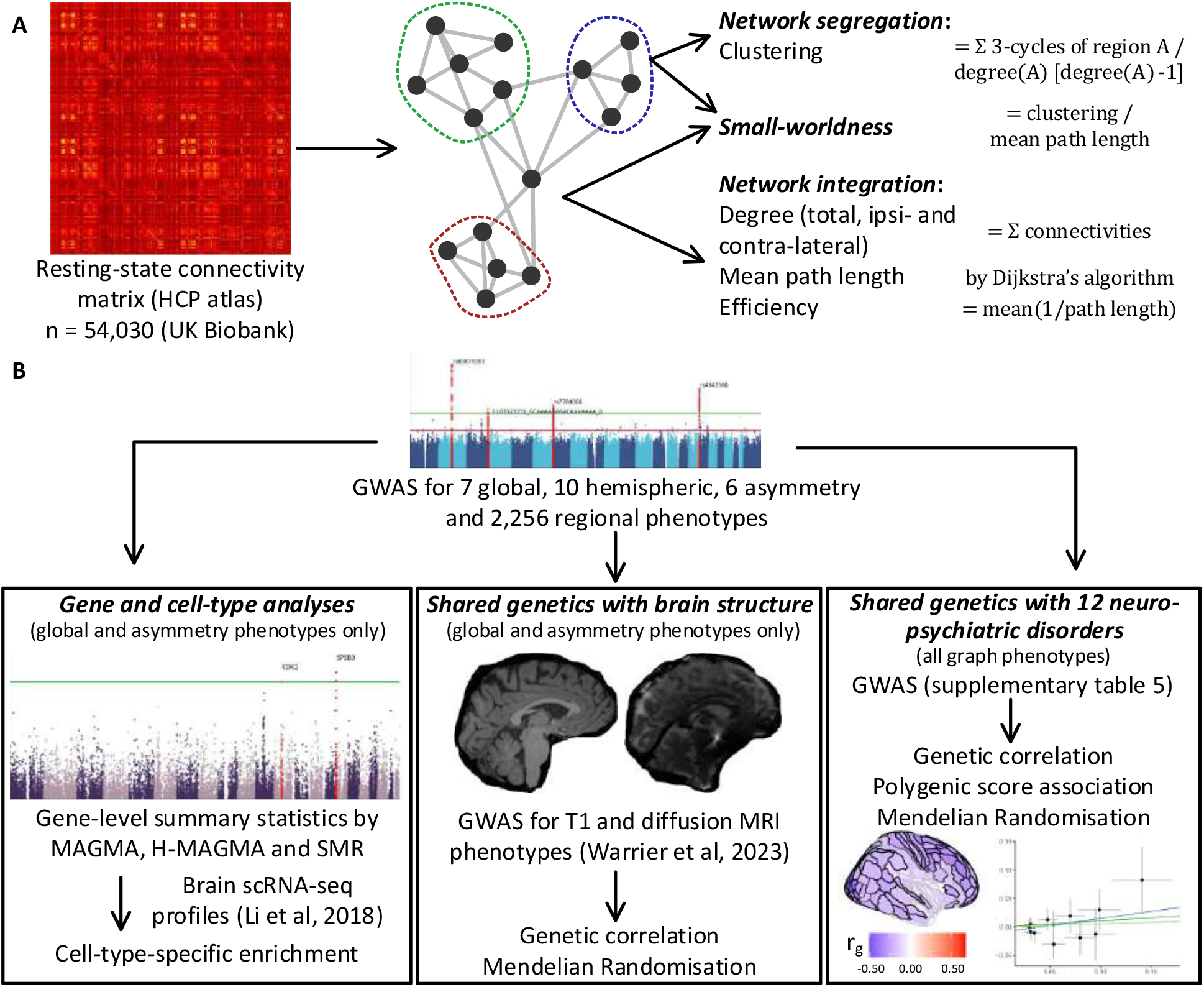
Schematic overview of the study. A. Schematic overview of the phenotyping protocol. Based on the 376-region connectivity matrix (left), we considered seven graph phenotypes: degree, the ipsi- and contra-lateral partitions of degree, efficiency, characteristic path length, clustering and small-worldness. The first five phenotypes measure network integration, clustering measures network segregation and small-worldness their balance. For hemispheric networks we omitted contralateral and total degree. Except for small-worldness, all other graph metrics were well-defined for individual brain regions and the correlation of corresponding regional phenotypes across left and right hemispheres gave the hemispheric asymmetry phenotypes. There were two additional parametrisations of asymmetry that were not further analysed due to low heritability. B. Schematic overview of the genetic analyses. We conducted GWAS over 7 global, 10 hemispheric, 6 asymmetry and 2,256 regional graph phenotypes using mixed linear modelling (Methods). On that basis, we identified candidate genes of global-level functional network topology using parallel methods including MAGMA, H-MAGMA and summary-data Mendelian Randomisation, and combined these results with cell-type-specific RNA sequencing data to identify cell-type enrichment of the graph phenotypes (left panel). To understand the overlapping genetics between global graph phenotypes, asymmetry and brain structure (middle panel), we used published GWAS summary statistics on brain macro- and microstructure to identify genetically correlations with the functional phenotypes, and explored any causal relationship with two-sample Mendelian Randomisation. The same methods were repeated to study the genetic overlap between graph phenotypes and 12 neuro-psychiatric disorders (right panel), and were reinforced by polygenic score-based associations. We also tested the spatial enrichment of disorder correlates by analysing the regional-level genetic correlations and polygenic score associations.

## 2 Results

### 2.1 Genome-wide associations of global network architecture

We first conducted GWAS for the seven global- and hemisphere-level graph metrics (global graph phenotypes) in the UK Biobank (n = 54,030, 25,604 male). These included five measures of network integration (degree, the ipsi- and contra-lateral partitions of degree, efficiency and characteristic path length), one measure of network segregation (clustering) and small-worldness which measures integration-segregation balance (Figure 1, Supplementary Figure 1) [37]. Network integration metrics quantify information transfer across distributed regions of the brain, assessed via the correlation of fMRI signals between regions. Conversely, segregation metrics capture the modular structure of densely interconnected local circuits that support specialised information processing, observed as high temporal coherence within functional communities [37]. Further descriptions of the phenotypes are provided in the Methods.

Across all GWAS, we identified one novel genome-wide significant (*p <* 5e-08) locus on chromosome 21 associated with clustering (sentinel single-nucleotide polymorphism/SNP rs16993684, p = 4.295e-08, Figure 2a-b). This SNP was nominally associated with all other global phenotypes (p = 5.176e-08 in ipsilateral degree to 1.231e-05 in path length, Supplementary Table 1.1, Supplementary Figure 2-3). We identified no significant association between this SNP and a range of other neuroimaging related phenotypes from previously published GWAS (Methods), suggesting relative specificity to graph phenotypes.

**Figure 2:**
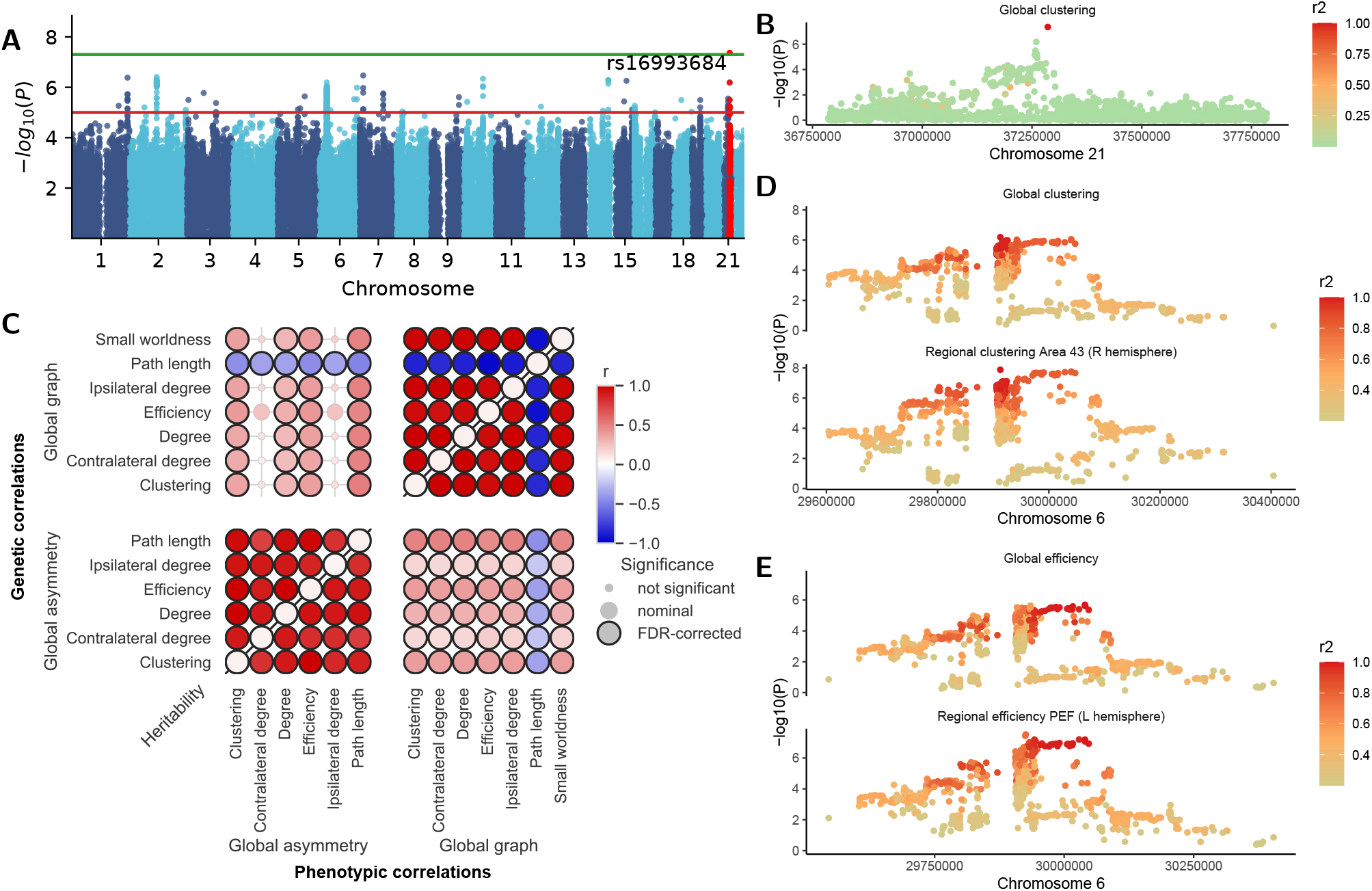
Genome-wide associations of the resting-state functional network topology. A. Manhattan plot for global clustering. Each point represents an SNP, and the Y axis is the significance level (p-value) on log_10_ scale. The green line is the genome-wide significance threshold (5e-08) and the read line is the suggestive threshold (1e-05). SNPs within *±* 1Mb from the top variant are marked red. GWAS was conducted using mixed linear modelling. B. Summary statistics zoomed in to *±* 0.5Mb from the significant variant (rs16993684) for global clustering. Colour scale represents the linkage disequilibrium (*r*^2^) with the top candidate SNP. C. Heritability (marked diagonal) and genetic correlations estimated by LDSC (above diagonal), and phenotypic Pearson correlations (below diagonal) between global graph metrics and hemispheric asymmetry of graph phenotypes. Colour scale indicates the correlation coefficient or heritability. Significant correlations after FDR correction are highlighted with dark borders. D-E. The genomic region in chrosome 6, 29.6-30.6Mb can be separated into two genetic clusters, with candidate SNPs being rs2735103 (D) and rs9393989 (E). Format is the same as (B).

Two previous studies have conducted GWAS of global graph phenotypes, albeit in smaller samples and using slightly different phenotyping protocols (Supplementary Note 1). Foo et al. [24] identified one locus (sentinel SNP rs62158161) associated with global network efficiency and characteristic path length, using a different parcellation (Supplementary Table 1.2). This locus was nominally significant in the present study (p = 6.16e-07 to 1.48e-04), suggesting that genetic associations of functional connectome topology is robust across parcellations. For another GWAS study on global efficiency based on only the top 10% of connectivity [23], we replicated four out of seven loci at nominal significance level (*p* < 0.05) in the present study. All global graph phenotypes showed low but significant SNP-based heritability (linkage disequilibrium score regression/LDSC-estimated *h*^2^ = 0.0558 ± 0.0094 to 0.0621 ± 0.0094, Supplementary Table 2.1).

Additionally, we also derived asymmetry for graph metrics using three different methods: absolute difference, fractional asymmetry score, and correlation of corresponding regions in both hemispheres (Methods, Supplementary Note 2). Only the correlation measure of asymmetry was heritable (LDSC *h*^2^ = 0.0486 ± 0.0086 to 0.0636 ± 0.0086, Supplementary Table 2.1) whilst the heritability of the other two methods was not significant (max *h*^2^ = 0.0038). Hence, for all subsequent analyses, we only used the correlation method for asymmetry phenotypes. We identified no genome-wide significant loci for the asymmetry phenotypes (Supplementary Figure 2). Top loci are given in Supplementary Table 1.1.

We assessed the replicability and generalisability of the global graph and asymmetry phenotypes using individuals of predominantly South Asian (N = 618) and African (N = 376) genetically-inferred ancestries in the UK Biobank who were not included in the original GWAS. The top SNP (rs16993684) was associated with clustering in the African-like population (p = 0.021, Supplementary Table 6.1), and in the consistent direction. Meta-analyses across the three populations increased the statistical significance of this association (European Discovery, p = 4.30e-08; Cross-ancestry meta-analysis, p = 3.22e-08). Furthermore, for all GWAS, meta-analysis of the global graph and asymmetry GWAS across the three populations increased the genomic inflation factor of the GWAS compared to the European-only discovery sample, as measured by LDSC (Supplementary table 6.2). This suggests largely consistent effect direction across the three populations. Because of the differences in linkage disequilibrium across populations, we restricted subsequent analyses to the European-only discovery GWAS.

To better understand the latent structure underlying different global graph and asymmetry phenotypes, we quantified the genetic and phenotypic correlation among all phenotypes (Figure 2c, Supplementary Figure 4, Supplementary Tables 2.2-2.3). As a sanity check, we confirmed that efficiency and path length were negatively correlated (|*r*_*g*_| = −0.970 ± 0.011, |*r*_*p*_| = −0.940) as expected by their mathematical definition (Methods). Whilst different graph metrics capture different aspects of network architecture [6, 7], the phenotypic and genetic correlations among all global graph phenotypes were high (|*r*_*p*_| ≥ 0.790, |*r*_*g*_| ≥ 0.814). All global graph phenotypes were positively correlated except for path length. All asymmetry phenotypes also had high phenotypic and genetic correlations (*r*_*p*_ ≥ 0.764, *r*_*g*_ ≥ 0.749). Global and asymmetry phenotypes were moderately correlated (|*r*_*p*_| = 0.139 to 0.491, |*r*_*g*_| = 0.073 to 0.519), all in the positive direction except for path length. Taken together, this suggests high shared genetics among global graph phenotypes and among asymmetry phenotypes, and moderate shared genetics between the two sets of phenotypes. The mirroring of the genetic and phenotypic correlations is consistent with Cheverud’s conjecture that phenotypic correlations are proxies for genetic correlations [43].

### 2.2 Genome-wide associations of regional graph phenotypes

To investigate common variant genetic influences on above graph metrics for each brain region (henceforth, regional graph phenotypes), we conducted GWAS for six graph metrics over 360 cortical regions of the Human Connectome Project parcellation atlas [38] and 16 sub-cortical regions (total 2,256 phenotypes). Because regional phenotypes were highly correlated with global graph phenotypes (Supplementary Figure 6), we did not adjust for global graph phenotypes, as adjusting for a heritable, correlated covariate leads to collider bias and increases false positive results [44, 45]. SNP-based heritability for regional graph phenotypes were lower compared to global graph phenotypes (*h*^2^ = 0.0086 ± 0.0093 to 0.0850 ± 0.0098, Supplementary Figure 5a, Supplementary Table 2.4), but 2,232 out of 2,256 (~ 99%) phenotypes were still significantly heritable. Across all regional phenotypes, we identified 175 loci at study-wide significance (*p* <3.11e-11, correcting for 1649.2 independent phenotypes, see Methods) and 1,194 at genome-wide significance. After accounting for overlapping loci between different brain regions, we identified seven study-wide significant and 180 genome-wide significant loci across six graph phenotypes. Amongst the 175 study-wide associations of the seven loci, 159 were available in other ancestries, of which 130 in the South Asian-like cohort and 105 in the African-like cohort were in consistent direction, and four were nominally significant (p *<* 0.05, Supplementary Table 6.3). Meta-analyses of the top SNPs across the three populations increased the statistical significance of 101 associations. Overall, this largely supports the replicability and generalisability of these loci across genetically-diverse ancestries. For regional asymmetry measured using fractional asymmetry score (Methods), we did not conduct GWAS as the SNP-based heritability was not statistically significant for most of the regions (max *h*^2^ = 0.034, 132/1128 nominally significant, Supplementary Figure 5b, Supplementary Table 2.5).

In order to understand the shared genetics across different brain regions, we conducted multi-trait colocalisation analysis [46]. This is a statistical approach that evaluates whether multiple traits share the same causal genetic variant at a genomic region (± 500 kb from sentinel variant) using a posterior regional association probability of *p* > 0.6 (Supplementary Table 1.4). We only tested genomic regions that contained at least one study-wide significant (*p* < 3.11e-11) SNP for regional graph phenotypes, or one SNP at significance level *p* < 5e-6 for global graph phenotypes or hemispheric asymmetry. The largest cluster (chromosome 6, 29.4-30.4 Mb) was shared across all global, hemispheric graph phenotypes and 119 to 201 brain regions for different regional phenotypes. The top two candidate SNPs in this region (rs2735103 and rs9393989, Figure 2d-e, Supplementary Figure 7) have both been associated with a range of immunological phenotypes [47], but no previous association has been reported for neuropsychiatric phenotypes. Another cluster on chromosome 16 (86.7-87.7 Mb) was associated with one asymmetry phenotype and path length of 13 brain regions (Supplementary Figure 8). The candidate SNP of colocalisation (rs12711472) is a known correlate of brain sulcal depth [48], subcortical volume [49], and in particular the volumes of 3rd, 4th and lateral ventricles [50], In addition, colocalisation also revealed 8 genetic clusters involving only regional phenotypes (Supplementary Table 1.4), but none of these clusters involved > 10 brain regions. Overall, these analyses suggest a complex genetic structure underlying global- and regional-level functional network organisation. Whilst the underlying genetics of many regional graph metrics overlap with global network phenotypes, hemispheric asymmetry also shows some overlapping genetic processes with regional network organisation.

### 2.3 Candidate genes for the functional connectome overlap with brain structure

To identify potential candidate genes underlying global-level network architecture, we prioritised candidate genes for global and asymmetry phenotypes using eleven methods (see Methods). Candidate genes were identified if they were supported by at least two parallel methods. In total, 10 genes were identified for global phenotypes (Figure 3a-b, Supplementary Figures 9-16, Supplementary Tables 1.5, 1.6, 4.1, 4.3). Four of these genes have been associated with structural development in the brain. Cyclin-D kinase 2 (*CDK2*), a gene critical for the G1/S phase of mitosis, is implicated in microcephaly, where multiple microcephaly-associated proteins form a complex and recruit *CDK2* to promote centrosome duplication [51]. It is also over-expressed in glioma [52]. *EGFLAM* has also been associated with glioblastoma by regulating cell cycle checkpoints [53]. *HLA-A* is involved in immune regulation and mutations have been implicated in immune tolerance to cancer and brain atrophy by inflammation [54–57]. *NUBP2* has been associated with neural crest organogenesis in mice although this has not been confirmed in humans [58]. No genes were consistently prioritised by multiple methods for asymmetry (Supplementary Tables 4.2, 4.4). Taken together, the association between these genes with both structural and functional brain phenotypes suggest the potential overlap of genetic processes underlying brain structure and function.

**Figure 3:**
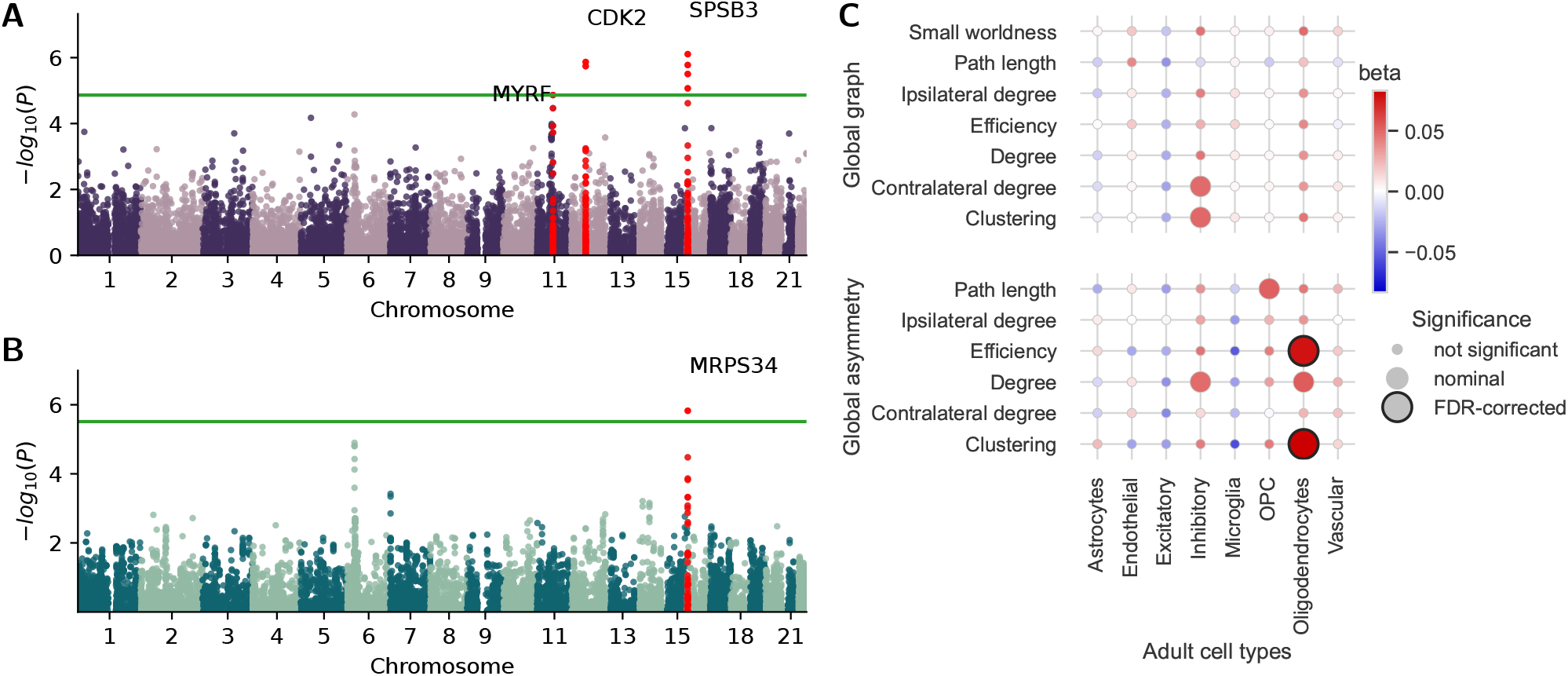
Gene and cell-type associations of the functional network. A. Aggregating SNP statistics by gene annotation using MAGMA identified 7 significant genes in 3 loci for small-worldness. B. Using summary-data Mendelian randomisation with splicing QTL identified *MRPS34* as a significant gene for small-worldness. For A and B, each point represents a gene and the y axis is the significance level (p-value) in log scale. The green line represents the genome-wide *p*_*fdr*_ = 0.05 threshold. Genes within ± 1Mb of each top gene are labelled red. C. Enrichment of global graph phenotypes and hemispheric asymmetry in different cell types in the adult brain. Colour scale represents the enrichment or depletion effect size estimated by MAGMA multiple regression with cell-type-specific gene sets in adult brains (Methods). Significant enrichments after FDR correction are highlighted with dark borders. OPC: oligodendrocyte precursor cells.

To understand the cellular processes associated with global functional networks, we investigated whether global and asymmetry phenotypes are enriched in different cell types in foetal and adult postmortem brain samples (Supplementary Tables 4.5-4.6). SNP-level summary statistics were aggregated to gene level using MAGMA [59] and tested for enrichment in post-mortem cell-type-specific expression datasets from adult and foetal brains [60]. We did not identify significant enrichment for global graph phenotypes but asymmetry was significantly enriched in oligodendrocytes in adults (Figure 3c).

### 2.4 Causal relationship between brain function and structure

The nature of the correspondence between the brain’s structural and functional connectivity has been an area of active debate. One theory suggests that functional connectivity is mainly constrained by the white matter connectivity backbone [29, 30]. Hence, we hypothesised that measures of microstructural integrity, which directly affect white matter connectivity [61], are also genetically correlated with functional connectivity. However, recent studies have demonstrated that the correspondence between microstructural connectivity and functional connectivity is imperfect [29, 30]. Another theory posits that the macroscale shape of the brain constrains brain function, thereby shaping macroscale functional connectivity of the brain [32]. We sought to test these two theories using genetic correlation and Mendelian randomisation. Given the heritability of both macro- and microstructural phenotypes [33] and functional networks, we hypothesized that any links between structure and function would be reflected in shared genetics, and causal relationship would be reflected in Mendelian randomisation analyses.

Global graph phenotypes and hemispheric asymmetry showed distinct phenotypic and genetic correlation patterns with global structural phenotypes, with genetic correlations mirroring phenotypic correlations (Figure 4a-b, Supplementary Figure 17, Supplementary Tables 2.6-2.7). Global path length was negatively correlated with global macrostructural phenotypes (*r*_*p*_ = −0.092 to −0.178, *r*_*g*_ = −0.156 to −0.254), including cortical volume, folding index and local gyrification index (a measure of the extent of the cortex buried within sulci), all of which are related to cortical expansion [33]. In parallel, we identified one colocalised genomic cluster between global graph phenotypes and cortical thickness (Supplementary Figure 18, Supplementary Table 1.7), but none between global graph and cortical expansion phenotypes, likely due to low statistical power. Genetic correlations with cortical microstructural phenotypes were low and not significant. Hemispheric asymmetry was most correlated with microstructure (isotropic volume, orientation dispersion index and mean diffusivity, *r*_*p*_ = 0.179 to 0.210, *r*_*g*_ = 0.265 to 0.518), consistent with the overlapping cell-type enrichment for both functional asymmetry (present study) and cortical microstructure [33] in adult oligodendrocytes. Supporting this genetic correlation, we identified co-localisation between all three microstructural phenotypes and functional asymmetry in chromosome band 16q24.2, which includes a known gene for hydrocephaly (*C16orf95*, Figure 4b, Supplementary Table 1.7) [62]. Taken together, we identified two cross-modality clusters between global graph phenotypes and cortical macrostructure, and between functional asymmetry and cortical microstructure.

**Figure 4:**
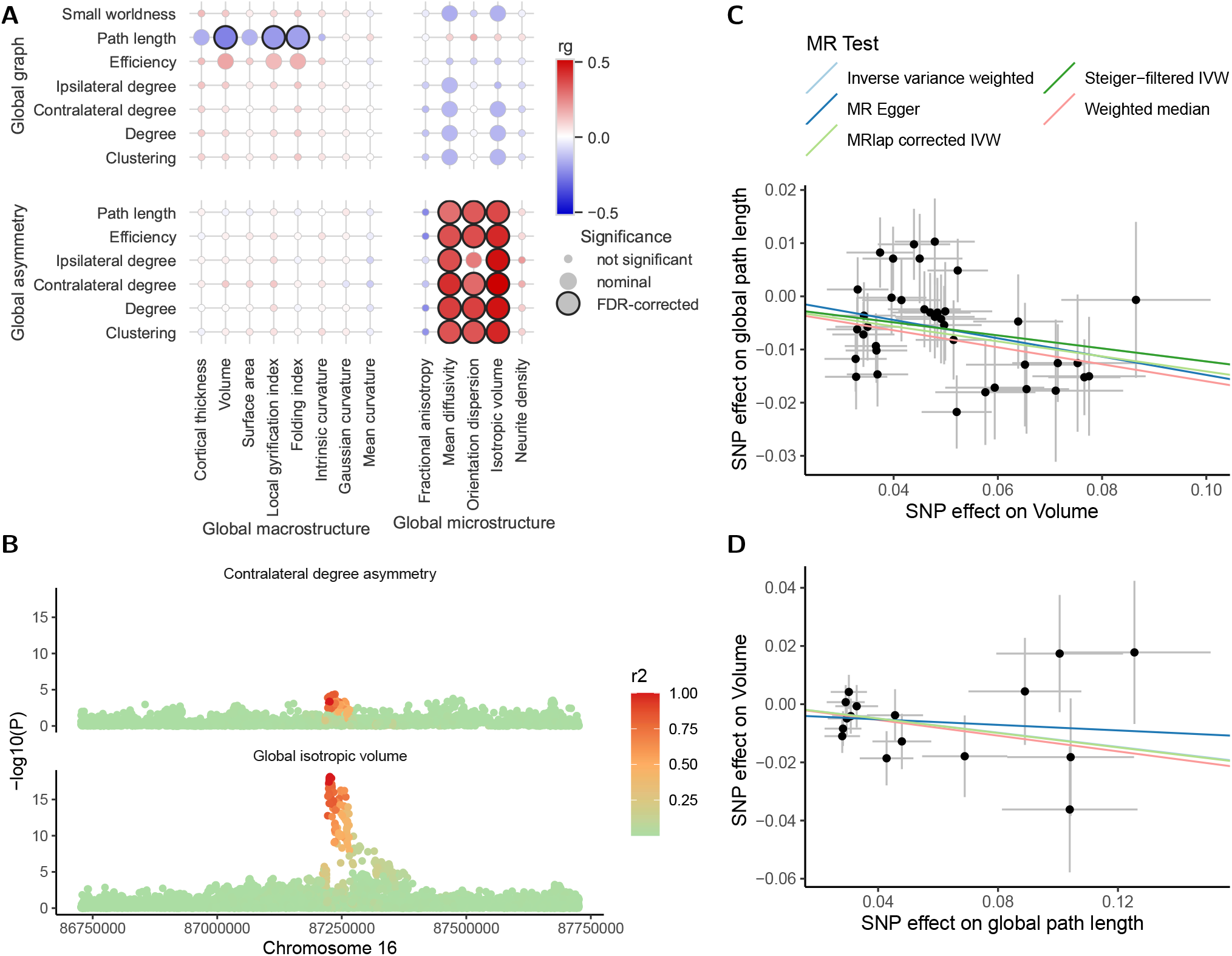
Shared genetics between graph phenotypes of the resting-state connectome, brain structure and functional gradients. A. Genetic correlation between global graph phenotypes, hemispheric asymmetry, and brain structure estimated using LDSC. Colour scale represents the genetic correlation coefficient and significant correlations after FDR correction are highlighted with dark borders. B. GWAS summary statistics zoomed in for the genomic region on chromosome 16 (86.7-87.7 Mb), where global isotropic volume was colocalised with hemispheric asymmetry. Each point represents a SNP, the y axis indicates the GWAS p-value in log scale and the colour scale represents its linkage disequilibrium with the candidate SNP (rs9933149). C-D. Mendelian randomisation scatter plot examining the causality from global volume to global path length (C) and from global path length to global volume (D), with estimated fits using five different methods. Each dot represents a SNP, and its x and y axis positions indicate its GWAS-estimated effect size on global path length and cortical volume. Error bars for each dot indicate its GWAS-estimated standard error. SNPs are only included in the analysis if they are significantly associated with the exposure, i.e. the phenotype assumed to be causally upstream to the other phenotype (see Methods).

Next, to assess if brain structure is indeed causally related to brain function, we conducted two-sample bidirectional Mendelian randomisation using MRlap, which accounts for sample overlap between GWAS studies. We confirmed that cortical volume (lowest MRlap *p*_*fdr*_ = 5.69e-04), folding index (lowest MRlap *p*_*fdr*_ = 1.16e-03) and local gyrification index (lowest MRlap *p*_*fdr*_ = 0.0250) were indeed causally related to global graph phenotypes (Figure 4c, Supplementary Figures 19-21). MRlap also identified a causal relationship between global cortical microstructure (isotropic volume and mean diffusivity) and functional asymmetry phenotypes (lowest MRlap *p*_*fdr*_ = 8.82e-11, Supplementary Figures 23-25). All results were supported by parallel Mendelian randomisation methods (Supplementary Tables 3.1-3.2). We found no evidence for causality in the reverse direction (Figure 4d). Overall, genetic variants associated with brain structure are causally related to brain function, but micro- and macro-scale structure affect different aspects of brain functional organisation.

We assessed if the shared genetics and causal relationship between cortical expansion and global graph phenotypes replicate in a different dataset, focussing on global path length which had the most significant genetic correlation with cortical expansion phenotypes. Whilst independent GWAS studies for volume, folding index and local gyrification index are not available, we chose surface area to replicate the above findings as it also shares the common latent genetic factor for cortical expansion [33]. We used GWAS summary statistics for surface area from the ENIGMA consortium [63], with partly overlapping participants (10,083 overlap) with the GWAS of cortical expansion phenotypes previously used in this study. We identified a significant negative genetic correlation between SA and global path length (rg = −0.218, p = 0.0021, Supplementary Table 6.4), and Mendelian Randomisation confirmed that surface area is causally related to global path length (Supplementary Table 6.4, Supplementary Figure 22).

Finally, we questioned if these findings are generalisable to other signatures of brain function. To test this, we obtained published GWAS results for functional gradients [64], and phenotypic data in the UK Biobank, and estimated the phenotypic and genetic correlations with graph phenotypes derived above (Supplementary Figure 17, Supplementary Tables 2.6-2.7). Whereas graph metrics of the functional connectome describe the small-world structure of the functional connectome and how this structure facilitates efficient information processing [7, 8, 37], diffusion-embedding based functional gradients describe the hierarchical organisation of information flow between brain regions [65, 66]. Functional gradients were quantified at global level using similarity to the group-based template in the Human Connectome Project [64]. The majority of the global graph phenotypes were moderately correlated with the second functional gradient (*r*_*p*_ = −0.133 to −0.199, *r*_*g*_ = −0.230 to −0.304), but their correlation with cortical macrostructure was not replicated in the second functional gradient. Conversely, asymmetry was more strongly correlated with all top three functional gradients (*r*_*p*_ = −0.140 to −0.282, *r*_*g*_ = −0.320 to −0.641), which were all correlated with cortical microstructural phenotypes [64]. These results support the generalisability of functional network topology (specifically asymmetry) to hierarchical gradients of functional networks, and suggest a more general applicability of structural constraints on functional organisation of the brain.

Taken together, these findings suggest that both hypotheses capture complementary aspects of brain structure-function relationships: macrostructural cortical shape appears to constrain global network topology and efficiency, while microstructural properties shape the asymmetric organization and hierarchical gradients of brain function. This dual constraint system may help explain how the brain achieves both local specialisation and global integration.

### 2.5 Genetic link between the functional connectome and autism

A substantial body of literature has linked neurodevelopmental and psychiatric conditions to alterations in global and regional functional connectivity patterns. For instance, one theory characterises autism as a condition with altered functional connectivity [67]. Evidence from MRI, magneto-, and electroencephalogram studies find support for altered functional connectivity in autism, although with substantial heterogeneity across studies [68]. Similarly, conditions like schizophrenia, depression, and attention-deficit hyperactivity disorder (ADHD) have all been variously linked to altered functional connectivity [69–71]. However, case-control differences in functional connectivity have long been linked with non-genetic factors such as duration and severity of the condition and medication use [72–75], so it is unknown to what extent the altered functional connectivity in neuropsychiatric conditions can be explained by shared underlying genetics. To investigate this, we estimated the genetic correlations between global and regional graph phenotypes of the functional network and 12 psychiatric and neurodevelopmental conditions (Figure 5a, Supplementary Figure 26, Supplementary Table 2.8). Global graph phenotypes were modestly but significantly genetically correlated with autism (*r*_*g*_ = 0.151 to 0.245) but none of the other conditions tested, suggesting relative specificity of the findings with autism. We replicated this finding by meta-analysing the genetic correlations obtained from two independent GWAS of autism [76, 77] and identified statistically similar genetic correlations (Supplementary Table 6.5). Further supporting this finding, polygenic scores for autism were associated with global graph phenotypes in 54,030 individuals in the UK Biobank in the consistent direction (Figure 5b, Supplementary Figure 27, Supplementary Table 2.9). We tested if these results are due to the shared genetics between global graph phenotypes and cortical structure, especially cortical expansion phenotypes. We found no significant correlation between cortical structure and autism (lowest nominal p = 0.051, Supplementary Table 2.8), suggesting that the shared genetics between functional graph phenotypes and autism is independent from cortical structure.

**Figure 5:**
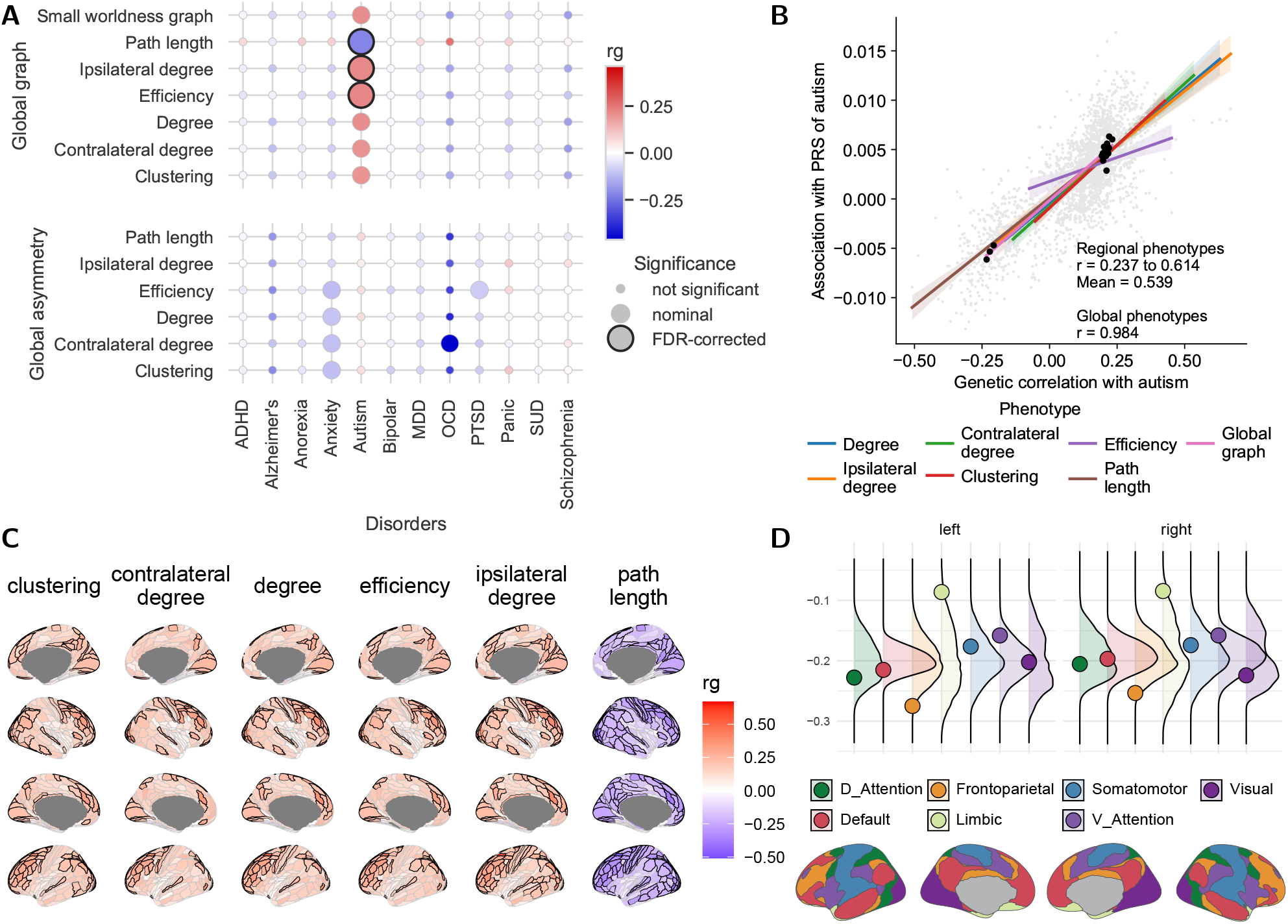
Shared genetics between the resting-state connectome and neuropsychiatric conditions. A. Genetic correlation between global graph phenotype**1**s, hemispheric asymmetry and 12 neuropsychiatric conditions, estimated with LDSC. Colour scale represents the genetic correlation coefficient and significant correlations after FDR correction are highlighted with dark borders. B. Genetic correlation (x axis) and polygenic score-based effect sizes (y axis) between different graph phenotypes and autism are highly similar, except for regional efficiency. Each pale dot represents one graph phenotype for one brain region. Dark dots represent global and hemispheric graph phenotypes. Lines are regression fits for different regional graph phenotypes across all brain regions, and the shading represents the regression error. C. Genetic correlation between regional graph phenotypes and autism estimated by LDSC. Significant correlations after FDR correction are highlighted with dark borders. D. For regional path length, spherical spin-permutation showed that frontoparietal regions were particularly correlated with autism. For each functional network of the Yeo-7 atlas, the coloured dot represents the mean genetic correlation with autism across all regions within that network, and the curve represents the spherically permuted null-distribution. The graphical legend indicates the spatial positioning of the Yeo-7 networks. ADHD: attention-deficit hyperactivity disorder. MDD: major depressive disorder. OCD: obsessive-compulsive disorder. PTSD: post-trauma stress disorder. SUD: substance use disorder.

To investigate if this association remains at a regional level, we quantified the genetic correlation and polygenic score-based association between regional phenotypes and autism. Overall, across the cortex, we identified moderate genetic correlation between regional phenotypes and autism in the consistent direction as the global graph phenotypes (*r*_*g*_ = −0.547 to 0.233 for path length and −0.331 to 0.702 for other phenotypes, Figure 5c, Supplementary Figure 28b, Supplementary Tables 2.10-2.11). We obtained consistent findings with polygenic scores for autism (Figure 5b, Supplementary Figure 28a, 28c, Supplementary Table 2.12). Spherical spin-permutation identified higher genetic correlations between autism and regional phenotypes in frontoparietal regions, where likelihood of autism increased with increasing regional connectivity (Figure 5d). This aligned with clinical findings that autistic individuals show hyper-connectivity in the frontoparietal network [78].

These results suggest moderate shared genetics between autism and both global and regional graph phenotypes. These findings are relatively specific to autism, and substantially larger in magnitude to the shared genetics between autism and cortical structural phenotypes [79]. Subsequently, to investigate a causal relationship between autism and the graph phenotypes, we conducted bidirectional two-sample Mendelian Randomisation. After correction for multiple testing, Mendelian randomisation analyses did not identify any significant causal effects between autism and global network phenotypes or vice versa (Supplementary Figure 29, Supplementary Table 3.3). This may be due to weak instruments used in the analyses or low statistical power.

Asymmetry phenotypes were nominally genetically correlated with a range of mood and anxiety disorders (anxiety, post-trauma stress disorder and obsessive-compulsive disorder, *r*_*g*_ = −0.052 to −0.463, nominal *p* < 0.05, Figure 5a, Supplementary Table 2.8). Association between asymmetry phenotypes and polygenic scores of these conditions were also in the consistent direction (Supplementary Table 2.9). Previous phenotypic studies have found altered functional asymmetry in ADHD and schizophrenia [26, 80]. In our analyses, however, the different measures of functional asymmetry were either not heritable, or genetically uncorrelated with ADHD and schizophrenia. This suggests that altered functional asymmetry observed in schizophrenia and ADHD are either sequelae of the two conditions and associated therapeutics, or due to shared environmental factors or rare genetic variants.

## 3 Discussion

In this study, we systematically analysed the common variant genetics of functional brain network topology using data from 54,030 individuals from the UK Biobank. We found graph metrics of functional networks to be modestly heritable, and identified one significant locus at global level and 11 at regional level (Figure 2, Supplementary Figures 2-7, Supplementary Tables 1.1-1.6), which we replicated in individuals of African-like and South-Asian-like genetic ancestries. Combined evidence from multiple methods identified 10 candidate genes for global graph metrics, including four genes previously associated with neurological disorders. On this basis, we were able to genetically validate several phenotypically-informed hypotheses about human brain function and its clinical relevance. Overall, our systematic genetic characterisation offered novel insights into the biology underlying human brain function.

Graph phenotypes have long been used to describe a small-world brain, which balances between segregation into highly connected modules and global-level integration [6–8, 10]. Converging evidence from imaging and molecular genetics suggested that segregation and integration are driven by short- and long-distance connections [81–83], and are associated with different domains of cognitive function [84]. However, results from this study suggested that individual differences in network segregation and integration are highly similar both phenotypically and genetically (Figure 2c, Supplementary Figure 4, Supplementary Tables 2.6-2.7). Regional network topology was also found to be genetically consistent with the global level. This suggests a potential overarching constraint over different aspects of functional network topology, which has been stipulated by the economic theory that functional network organisation is a trade-off between processing efficiency and energy costs in network wiring and maintenance [10, 85].

We found replicable evidence of overlapping genetics between cortical structural expansion and functional network topology (Figure 4, Supplementary Figures 17-25, Supplementary Tables 1.7, 2.6-2.7, 3.1-3.2), providing one explanation for concurrent changes of functional network architecture and cortical expansion at both developmental and evolutionary time-scales [83, 86, 87]. The causal link from cortical expansion to brain function supported the theory of geometric constraint on brain function [32]. Initial evidence suggested spatial overlap between evolutionary cortical expansion and structural connectivity hubs with wide-ranging long-distance neuronal connections [88–90]. It is possible that biological processes involved in cortical expansion promote the formation of long-range neuronal connections, providing the backbone for increased functional network efficiency. Conversely, for asymmetry and gradient hierarchy of the functional network, we observed shared genetics with cortical microstructure. Although cortical microstructure is not a measure of structural connectivity *per se*, this points to the role of microstructural integrity in shaping regional differences in functional connectivity. The combined results suggest that the architecture of the brain functional network is subject to dual constraints by cortical macro- and microstructure.

Although graph and asymmetry metrics of the functional network have been phenotypically associated with many neuropsychiatric conditions [14–17], we found minimal evidence for corresponding genetic correlations except for autism (Figure 5, Supplementary Figures 26-29, Supplementary Tables 2.8-2.12, 3.3). One explanation for this discrepancy is that the observed differences in functional connectivity are sequelae of neuropsychiatric conditions including duration and severity of the condition and medication use [72–75]. An alternate explanation is that there are shared non-genetic factors that contribute to both changes in functional connectivity as well as neuropsychiatric conditions [91, 92]. Further clarification is needed to ascertain if graph metrics are indeed endophenotypes for neuropsychiatric conditions.

Autism was a notable exception in our analyses. We observed robust and replicable shared genetics between autism and several graph phenotypes, and this was enriched in frontoparietal regions. A rich body of literature has investigated connectivity differences in autism, with substantial variation in the literature [93]. Studies have characterised autism as a condition marked by both hyper- and hypoconnectivity, with some studies identifying functional atypicalities in the frontoparietal network [78]. Taken together with this work, our findings provide orthogonal evidence for the overlap between autism and differences in functional connectivity. However, it is clear that autism is highly heterogeneous condition, with variation in polygenic profiles [94, 95]. Future analyses should investigate if the observed association differs along different axes of heterogeneity in autism.

Our analyses focused mainly on individuals of predominantly European ancestry, because for each other ancestry group, there were only 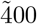 individuals which did not offer sufficient statistical power for GWAS by mixed linear modelling. However, further analyses suggests that above findings are largely replicable across different genetic ancestries. We also only conducted analysis in older adults (UK Biobank only recruits participants *≥* 40 years of age). Whilst realising that functional network organisation is a non-linear dynamic process throughout the human lifespan [96], we also did not have comparable statistical power in younger cohorts, due to sample size and imaging quality. The expansion of imaging genetics datasets across different age groups and ethnicities will paint a more complete picture of genetic effects on the development of functional and structural organisation of the human brain. We also recognise the heterogeneity of methods for image pre-processing and phenotype derivation, for example in asymmetry scoring [39, 40, 97, 98], connectivity parametrisation [99] and parcellation atlases [100]. Our study provided initial evidence for the generalisability of genetic findings across functional brain phenotypes. By making our results public, we hope further studies using parallel approaches will further consolidate this notion.

In conclusion, we provided novel insights into the genetic underpinnings of functional network architecture, their overlaps with brain structure and relevance to neuropsychiatric conditions. This work complements previous systematic studies [33, 101] in creating a cross-modality genetic atlas of imaging phenotypes which will benefit further analyses.

## 4 Methods

### 4.1 Phenotyping

#### 4.1.1 Image pre-processing

We used UK Biobank as our study cohort. UK Biobank is a large-scale health database including around 500,000 participants with whole-genome and microarray genetic data, metabolic biomarkers, questionnaires and digital health records from the UK National Health Service. Brain images of different modalities have been acquired for around 60,000 individuals. De-noised resting-state functional MRI images for 56,627 individuals were obtained from the UK Biobank (490 volumes TR/TE = 735/39 ms, multiband factor 8, voxel size 2.4 × 2.4 × 2.4 mm, FA = 52 degrees, FOV 210 × 210 mm). Pre-processed images were then parcelled according to the Human Connectome Project into 360 cortical and 16 sub-cortical regions [38] by performing a surface-based alignment of the atlas to each individual’s structural T1 and T2-FLAIR images (pre-processed as in [102]). FSL FLIRT [103] was used for functional-structural alignment. For each individual, time series of each region were filtered for wavelets using a Daubechies filter with four moments, cross-correlated using Pearson correlation, and Fisher transformed to z-scores to obtain the resting state connectome. Self-connections and negative connections were set to zero and each connectome was standardised to the range [0,1] as required by the Brain Connectivity Toolbox [37] (Supplementary Note 1). We chose the Pearson correlation to parametrise connectivity edges for multiple reasons: From the imaging perspective, compared to partial correlation and information-based parametrisations, Pearson correlation is most sensitive to the community structure between brain regions, which is subsequently described by graph metrics. Pearson correlation also has better test-retest reliability and preserves inter-individual differences better than partial correlation [99]. From the genetic perspective, partial correlation adjusts for heritable, correlated effects, which will bias the GWAS effect estimates [44]. For these reasons, we used full correlation to parametrise the functional connectivity matrix.

#### 4.1.2 Graph phenotype derivation

Following graph theory metrics for each region and for the global network were calculated using the Brain Connectivity Toolbox [37]:

1. Degree: the sum of connection weights between a given brain region and every other region. For connectivity matrix *A*, regional degree *d*_*i*_ = _*j*_ *A*_*ij*_, global/hemispheric degree *D* = ∑_*i*_ *d*_*i*_/2 = ∑_*i* ≠ *j*_ *A*_*ij*_.

2. Contralateral degree: the sum of connection weights between a given brain region and all regions of the opposite hemisphere. Same formula as above except that only brain regions in the opposite hemisphere are counted.

3. Ipsilateral degree: the sum of connection weights between a given brain regions and other other regions of the same hemisphere. Same formula as above except that only brain regions in the same hemisphere are counted.

4. Clustering coefficient: measures the density of connections amongst the neighbours of each brain region, where neighbours means regions connected to the given brain region. For weighted networks, clustering coefficient is the geometric mean of all triangles associated with each node, i.e. regional clustering *c*_*i*_ = mean_*j,k*_ (*A*_*ij*_*A*_*jk*_*A*_*ki*_)^1*/*3^, global/hemispheric clustering *C* = mean_*i*_(*c*_*i*_).

5. Characteristic path length: the arithmetic mean of the shortest path length between a given brain region and every other region, *L*_*ij*_, calculated by the Dijkstra’s algorithm. Regional path length *λ*_*i*_ = mean_*j*_(*L*_*ij*_), global/hemispheric path length Λ = mean_*i,j*_(*L*_*ij*_).

6. Efficiency: regional efficiency for each brain region is the harmonic mean of path length to all its neighbours, i.e. *e*_*i*_ = mean_*i,j*_(1*/L*_*ij*_); global and hemispheric efficiency is the harmonic mean of all path lengths within the global or hemispheric network, i.e. *E* = mean_*i,j*_(1*/L*_*ij*_).

7. Small-worldness: clustering coefficient divided by characteristic path length of a network,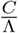. It is defined for global and hemispheric networks only.

We calculated 1-6 for each of the 376 regions (regional phenotypes), 3-7 for each hemispheric network (hemispheric phenotypes) and 1-7 for the global brain network (global phenotypes). All phenotypes above were linearly scaled such that the variance across the subjects is 1.

Based on the regional phenotypes, we further derived the hemispheric asymmetry of metrics 1-6 using a rank-based method previously used to analyse the structural connectome [39, 40]. Taking efficiency as an example, we calculated the Spearman correlation coefficient between the local efficiencies of the 188 brain regions in the left hemisphere and those of the right hemisphere, and converted the result into z-values by Fisher transformation. To ensure higher z-scores reflect higher asymmetry, the correlative asymmetry score was multiplied by −1, i.e.

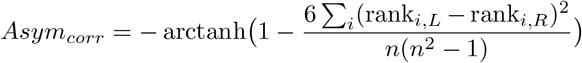

where *n* = 188, and *i* stands for each brain region. Using hemispheric phenotypes above, we also calculated the fractional asymmetry score [97, 98]

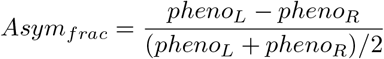

and the absolute difference (L - R) as candidate measures of asymmetry, but because these measures were not heritable, only the first correlative measure was included in downstream analyses. Finally, we also derived regional-level asymmetry using the fractional asymmetry score but these were also not included due to low heritability.

### 4.2 Genome-wide association study (GWAS)

#### 4.2.1 Genetic quality control

We used the previously quality controlled genotype array and imputed data provided by the UK Biobank [104]. Genomic data were quality-controlled by including people with self-identified European ethnicity. From this group, we excluded individuals above *±* 5 standard deviations from the mean in the first two genetic components, with genotyping rate *<* 95%, with excessive genetic heterozygosity, and those with non-matching genetic and self-reported sex. We also excluded individuals with poor imaging quality by filtering for mean framewise displacement, max framewise displacement and Euler index being within 5 median absolute divergence from population median, resulting in 56627 individuals. We then filtered the variants on minor allele frequency > 1%, call rate > 95%, not violating Hardy-Weinberg equilibrium (*p* > 1 × 10^−6^) and, for imputed SNPs, on imputation *r*^2^ > 0.4. After quality control and merging genetic and imaging datasets, we included a total of 9,142,950 SNPs for 54,030 individuals in our analysis. We did not consider other ethnicity groups because there were < 400 individuals for each ethnic group with the full genotypic and phenotypic data, which does not offer sufficient statistical power for GWAS with mixed effect linear models (MLM).

#### 4.2.2 Discovery GWAS

We constructed a genetic relatedness matrix using all SNPs included in the GWAS with GCTA, and then used fastGWA-mlm [105] to conduct 2,279 GWAS for 7 global, 10 hemispheric, 6 asymmetry and 2,256 regional phenotypes. For all GWAS, we regressed out age, age^2^, sex, age × sex, age^2^ × sex, site of image acquisition, the first 40 genetic principal components, mean and maximum frame-wise displacement, and Euler index [106] as covariates. For regional phenotypes, we did not include the corresponding global graph phenotypes as covariates because regressing out a correlated, heritable trait would introduce a bias to the GWAS results [44, 45]. Since loci with effect in multiple regions may be eliminated, controlling for global phenotypes may also bias down-stream cross-regional comparisons. By matrix decomposition we identified 1650.55 independent factors and hence the experiment-wide significance threshold was set at 5e-8 / 1649.2 = 3.110e-11 [107]. To identify significant loci, we clumped the GWAS summary statistics using PLINK 1.9 [108] at a *r*^2^ threshold over 1000 kb, using LD information from all the individuals included in the current analysis. To investigate associations of significant loci with other phenotypes from published GWAS studies, we conducted phenome-wide association study on three different platforms [109–111].

#### 4.2.3 Replication GWAS

We investigated whether the findings were replicable and generalisable to different genetically-inferred ancestry populations of the UK Biobank. We focussed on the two largest groups of non-European genetically inferred ancestries: South Asian-like (N = 618), and African-like (N = 376). Same genomic and phenotypic quality control procedures were applied to these individuals. We used GENESIS [112] to conduct GWAS by mixed linear modelling using the same covariates as the discovery GWAS. We used a different method because the replication sample size was considerably smaller and GENESIS has been shown to have higher statistical power compared to fastGWA, at the expense of higher computational demand [113]. For global graph phenotypes and asymmetry, we conducted GWAS over all SNPs included in the discovery GWAS, and meta-analysed the results using inverse variance weighting by PLINK 1.9 [108]. We then compared the genomic inflation factor (lambda) as estimated by LDSC [114, 115] between the meta-analysed and discovery GWAS, where an increase in lambda indicates consistent association direction between discovery and replication samples. For regional graph phenotypes, we only conducted GWAS and meta-analysis for study-wide significant SNPs (*p* < 3.110e-11) due to computational resource constraints.

#### 4.2.4 Heritability analysis

We used linkage disequilibrium score regression (LDSC) [114, 115] to assess the SNP-based heritability for all phenotypes, using default settings and pre-calculated LD reference panels from the 1000 Genomes Project phase 3 release. Additionally, because LDSC might introduce a downward bias in the heritability estimate, we used GCTA-GREML [116] to obtain a parallel estimate of heritability for global and asymmetry phenotypes, using the same individuals, genotypes and covariates as in the GWAS study, and the genetic relatedness matrix as derived above. However, due to the large computational demand of GCTA-GREML, it was not used for regional phenotypes.

### 4.3 Correlation and causality analysis

#### 4.3.1 Genetic correlation

We obtained summary statistics for 13 global structural brain phenotypes [33], surface area from the ENIGMA consortium for replication analyses [63], 6 phenotypes for global gradient similarity with the Human Connectome Project reference [64], and 12 neuropsychiatric conditions (Supplementary Table 5) [117–127]. For autism, we used the latest, unpublished GWAS from iPSYCH which consists of 19,870 autistic individuals (15,025 males) and 39,078 controls (19,763 males). We used this GWAS because it had a larger sample size and better statistical power compared to the latest publicly available GWAS (2019) for autism (mean *χ*^2^ = 1.23 for the unpublished GWAS and 1.2 for the 2019 GWAS) [128]. We replicated these findings using independent GWAS from SPARK [77] and PGC[76], where we conducted separate genetic correlations and meta-analysed the estimates using inverse variance weighted meta-analysis. Genetic correlation between graph phenotypes and these phenotypes were estimated with LDSC using the same setup as above. Phenotypes that significantly correlate at global levels were further investigated for regional genetic correlations. For regional graph phenotypes, we further tested for enrichment of genetic correlation within functional networks [4] using 10,000 spherical spin-permutations as a null model [129].

#### 4.3.2 Colocalisation

HyPrColoc [46] was used to identify colocalised genomic regions across multiple phenotypes. Candidate genomic regions were selected based on study-wide significant loci (*p* < 3.1076e-11) for regional phenotypes, genome-wide significant loci (*p* < 5e-08) for global structural phenotypes and, because few loci were identified, at a relaxed threshold (*p* < 5e-6) for graph phenotypes and functional gradients. Genomic regions up to 500 kb up- and down-stream from these loci were analysed using the branch and bound divisive clustering algorithm incorporated in the HyPrColoc package, in order to identify clusters of colocalised traits. We used the default prior probabilities, i.e. prior.1 = 1e-4 (prior probability of a SNP being associated with one trait), prior.c = 0.02 (prior probability that an additional trait is associated with a SNP, given that SNP is associated with one trait). Colocalised genetic clusters of traits were identified at a regional association probability threshold of 0.6, because the empirical statistical power at this threshold was *>* 90% as shown in simulations by the original authors [46]. Colocalisation of global and asymmetry phenotypes with (a) regional phenotypes, (b) structural and gradient phenotypes and (c) disorders were analysed separately.

#### 4.3.3 Mendelian randomisation

Mendelian Randomisation is a statistical technique that utilises genetic variation to infer causal relations between phenotypes, comparable to a randomised controlled trial where individuals are randomised to different genetic variants [34]. To investigate the causal effect between global, asymmetry, structural brain phenotypes and neuropsychiatric conditions, we conducted Mendelian Randomisation using several parallel methods. We only performed Mendelian Randomisation where (a) significant genetic correlation was identified, and (b) a clear biological hypothesis can be formulated. To ensure there are > 5 genetic instruments for each phenotype, genetic instruments were selected at a p-value threshold of 5e-06 for global and asymmetry graph phenotypes and autism, 5e-08 for global structural phenotypes. Summary statistics were clumped at these thresholds using a LD window of 1 Mb to identify independent loci as instrumental variables. We then used MR-lap [130] as our primary method because it corrects for sample overlap and adjusts for winner’s curse in genetic instrument selection. In parallel, we also conducted MR-PRESSO to correct for outliers [131], MR-Egger and MR by weighted median to account for invalid instruments, and the traditional inverse variance weighted MR (MR-IVW). MR-Egger, MR by weighted median and MR-IVW were conducted using the TwoSampleMR R package [132]. Causal relationships between traits were identified if following criteria were satisfied:

1. MR-lap, MR by weighted median and MR-IVW were all significant at *p*_*fdr*_ < 0.05.

2. If outliers were identified by MR-PRESSO, the outlier-corrected p-value was also significant at *p*_*fdr*_ < 0.05.

3. MR-Egger shows a consistent effect direction with all the above Mendelian randomisation methods (because MR-Egger has lower statistical power, we did not require it to be significant).

4. If Steiger directionality test [133] showed incorrect effect direction, above analyses were repeated after filtering out variants that had a larger effect size on the outcome phenotype than the exposure phenotype. If these repeated analyses were still significant, a significant causal relationship was identified.

#### 4.3.4 Polygenic risk score-based correlations

Polygenic scores were generated for the 12 disorders using summary statistics excluding the UK Biobank. We used polygenic score by continuous shrinkage (PRS-cs), a Bayesian algorithm that infers posterior effect sizes without requiring p-value thresholds [134]. Default parameters were used (Gamma priors a = 1, b = 0.5, shrinkage factor *ϕ* = 0.01). Polygenic scores were then estimated based on these effect sizes, and all common (minor allele frequency > 0.1%) SNPs that are present in the both the UK Biobank and the HapMap3 European-ancestry reference panels. Resultant polygenic scores for each individual were then correlated with graph phenotypes correcting for the same covariates as used in the GWAS. Regional polygenic-score based associations were tested for spatial enrichment using the spherical spin permutation method described above.

### 4.4 Gene prioritisation and enrichment analysis

#### 4.4.1 Gene prioritisation

Gene-based analysis was done for global and asymmetry phenotypes using several parallel methods. (1) We used MAGMA (v1.10) [59] to aggregate SNP-level summary statistics to genes based on genomic position. A conservative flank size of 35 kb upstream and 10 kb downstream was used to capture regulatory elements for each gene, whilst reducing noise. In addition, we also used H-MAGMA [135] to map SNPs onto genes based on long-range chromatin accessibility regulatory interactions. We used Hi-C data from (2) foetal brain [136], (3) adult brain [137], and two cell-type-specific Hi-C datasets from (4) midbrain dopaminergic neurons [138], and (5) cortical neurons [139]. In parallel, we used (6) expression, (7) isoform and (8) splicing quantitative trait loci data from the PsychEncode Capstone II release [140] and conducted summary-data Mendelian randomisation (SMR) [141]. Significant genes, isoforms and transcripts were prioritised if they reach *p*_*fdr*_ < 0.05 within each genome. We excluded genes where heterogeneity in dependent instruments (HEIDI) test suggested that the significant SMR result was due to pleiotropy (HEIDI *p* < 0.01).

(9) In parallel, we conducted Bayesian inference-based functionally informed fine-mapping using Polyfun [142] on all loci with *p* < 5*e* −6 over a range of 500 kb up- and down-stream, to identify potentially causal variants. We included loci that fine-mapped to *<* 5 credible variants or less. (10) We also included candidate variants from colocalisation analyses where global and asymmetry phenotypes colocalised with regional phenotypes, if the candidate variant explained > 95% of the posterior regional association probability. For these variants, we used Ensembl Variant Effect Predictor [143] to investigate the functional annotations of potentially causal variants. (11) Finally, we prioritised the closest genes to sentinel variants of each loci. Genes were prioritised if at least two parallel methods supported the same gene.

#### 4.4.2 Enrichment analyses

For global and asymmetry phenotypes, we tested for their enrichment in different cortical cell types in foetal and adult samples. To construct cell-type-specific gene sets, we used ATAC data from foetal [144] and postnatal cortical cells [145], log-transformed and normalised gene expression values within each cell type, divided by the cross-cell-type mean gene expression values to get the relative expression value for each gene, and selected genes with the top 10% relative expression value for each cell type. We then tested gene-level summary statistics from above for enrichment in different gene sets using MAGMA [59]. Significant gene sets were identified after Benjamini-Hochberg correction.

## Ethics statement

UK Biobank has approval from the North West Multi-centre Research Ethics Committee (MREC) as a Research Tissue Bank (RTB) approval. This approval means that researchers do not require separate ethical approval for accessing and analysing de-identified data as was done in this study, and can operate under the RTB approval.

## Data availability

All GWAS summary statistics for graph phenotypes will be made publicly accessible on University of Cambridge servers upon publication. Summary statistics for autism can be requested from Jakob Grove. Summary statistics for other neuropsychiatric conditions have been made publicly available through the relevant articles listed in Supplementary Table 5. Please refer to the data availability sections of the respective articles.

## Code availability

We used standard statistical genetics packages for this study. These include fastGWA and GCTA-GREML (https://yanglab.westlake.edu.cn/software/gcta/#Overview), LDSC (https://github.com/bulik/ldsc), hyperColoc (https://github.com/cnfoley/hyprcoloc), Two-sample MR (https://mrcieu.github.io/TwoSampleMR). Code for phenotyping is available at https://github.com/yh464/2023-rsfc_gwas; code for genetic quality control is available at https://github.com/yh464/genetics_qc; code for GWAS and down-stream analysis is available at https://github.com/yh464/gwas_pipeline.

## Acknowledgements

This research was supported by funding from the Simons Foundation for Autism Research Initiative, the Well-come Trust (214322 *\* Z *\*18 *\*Z), Horizon-Europe R2D2-MH (grant agreement number 101057385), and UKRI (10063472), and the ImmunoMIND hub as part of the UKRI Mental Health Platform. For the purpose of open access, we have applied a CC BY public copyright licence to any author-accepted manuscript version arising from this submission. S.B.-C. also received funding from the Autism Centre of Excellence, the Templeton World Charitable Fund, the MRC and the National Institute for Health Research Cambridge Biomedical Research Centre. All research at the Department of Psychiatry in the University of Cambridge is supported by the NIHR Cambridge Biomedical Research Centre (NIHR203312) and the NIHR Applied Research Collaboration East of England. The views expressed are those of the author(s) and not necessarily those of the NIHR or the Department. Some of the results leading to this publication have received funding from the Innovative Medicines Initiative 2 Joint Undertaking under grant agreement no. 777394 for the project AIMS-2-TRIALS. This joint undertaking receives support from the European Union’s Horizon 2020 research and innovation program and the EFPIA and Autism Speaks, Autistica and the SFARI. A.D.B and J.G. was supported by grants from the Lundbeck Foundation (R102-A9118, R155-2014-1724, and R248-2017-2003). High-performance computer capacity for handling and statistical analysis of iPSYCH data on the GenomeDK HPC facility was provided by the Center for Genomics and Personalized Medicine and the Centre for Integrative Sequencing, iSEQ, Aarhus University, Denmark (grant to A.D.B.). R.R-G was funded by the EMERGIA Junta de Andalucía program (EMER-GIA20 00139), the Plan de Consolidación (CNS2023-14364) and the Plan de Generación de Conocimiento from the Agencia Estatal de Investigación (PID2021-122853OA-I00). S.L.V. was supported by the Max Planck Society through the Otto Hahn Award; the Helmholtz International BigBrain Analytics and Learning Laboratory (Hiball); the Jacobs foundation research fellowship; Hector foundation research development award.

