## Supplementary Figures and Notes for "GWAS of macro-scale resting state functional brain networks identify shared biology with brain structure and autism"

June 4, 2025

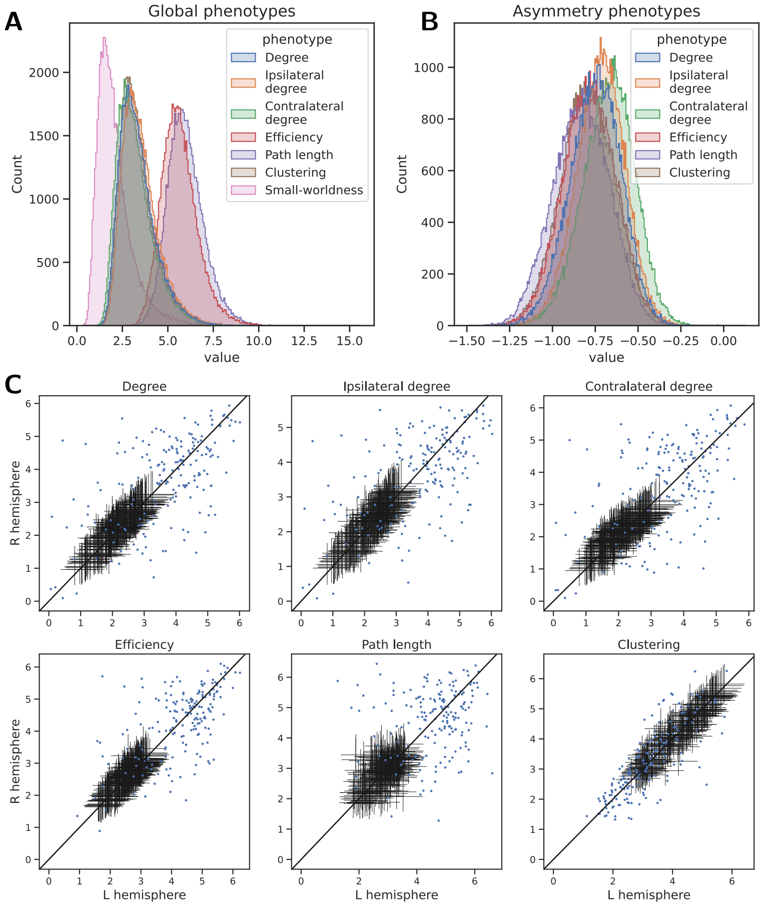


Supplementary Figure 1: **Distribution of the graph phenotypes included in this study.** A. Histogram for the seven global graph phenotypes. B. Histogram for the six hemispheric asymmetry phenotypes measured using the correlative measure. C. Distribution of the regional graph phenotypes. Each sub-plot depicts one graph phenotype for all 376 brain regions. Black error bars show the median and quartiles of each region in the left hemisphere (X axis) and its corresponding region in the right hemisphere (Y axis) across the 54,030 individuals. Blue points show the regional graph phenotype values for a random individual as an example.

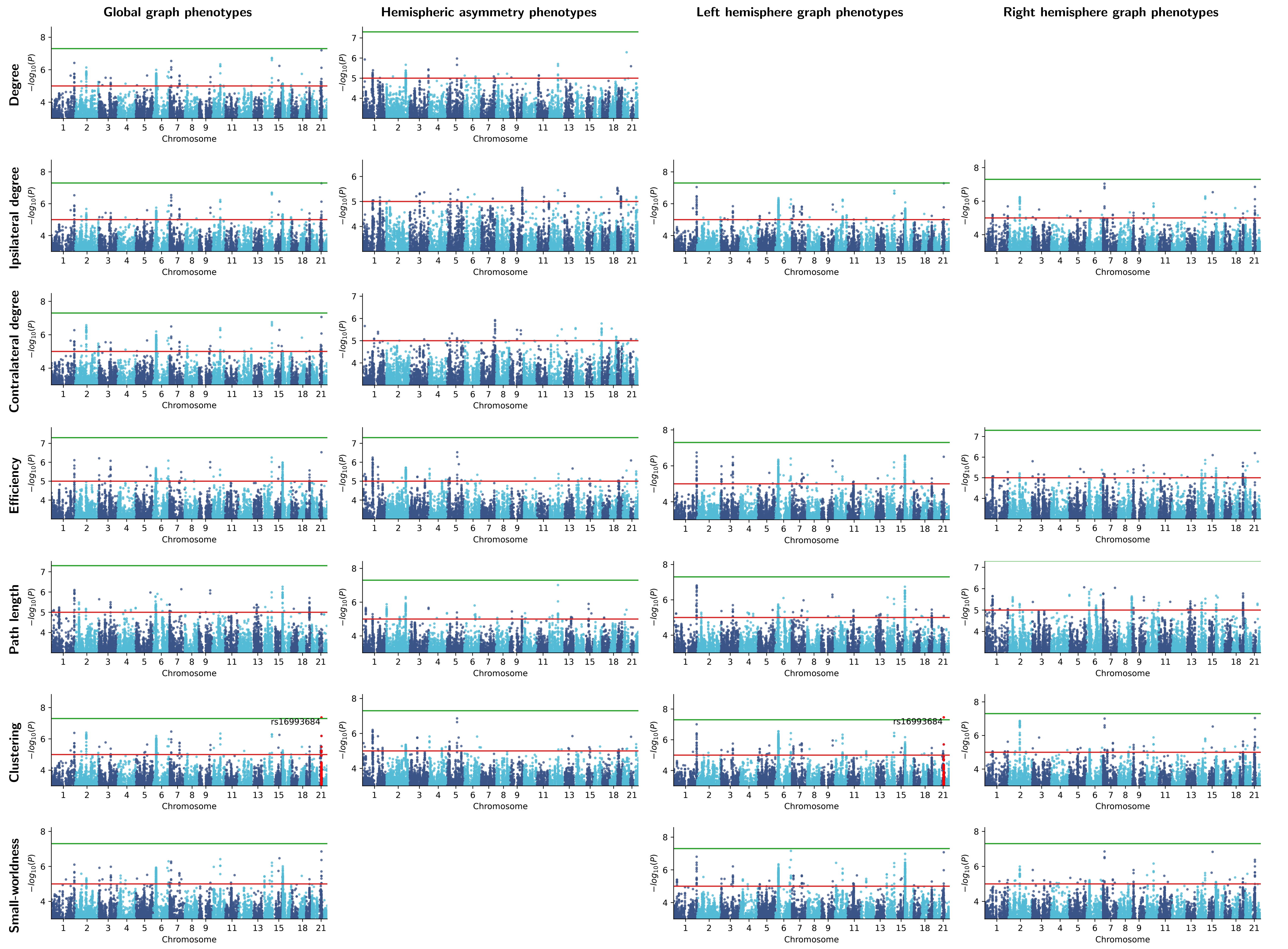


Supplementary Figure 2: **Manhattan plots for GWAS association results across all global, hemispheric and asymmetry phenotypes.** Each point represents a single nucleotide polymorphism (SNP), the X axis position represents its genetic position and the Y axis its p-value in log scale. The green line is the genome-wide significance threshold (5e-08). The red line is the suggestive threshold (1e-05). All significant SNPs and SNPs within ± 1Mb are labelled red. The IDs of the top SNP for each locus are labelled. Only SNPs at significance level $p < 0.001$ are included.

Supplementary Figure 3: **Manhattan Q-Q plots for all global, hemispheric and asymmetry phenotypes.** Each point represents a SNP.

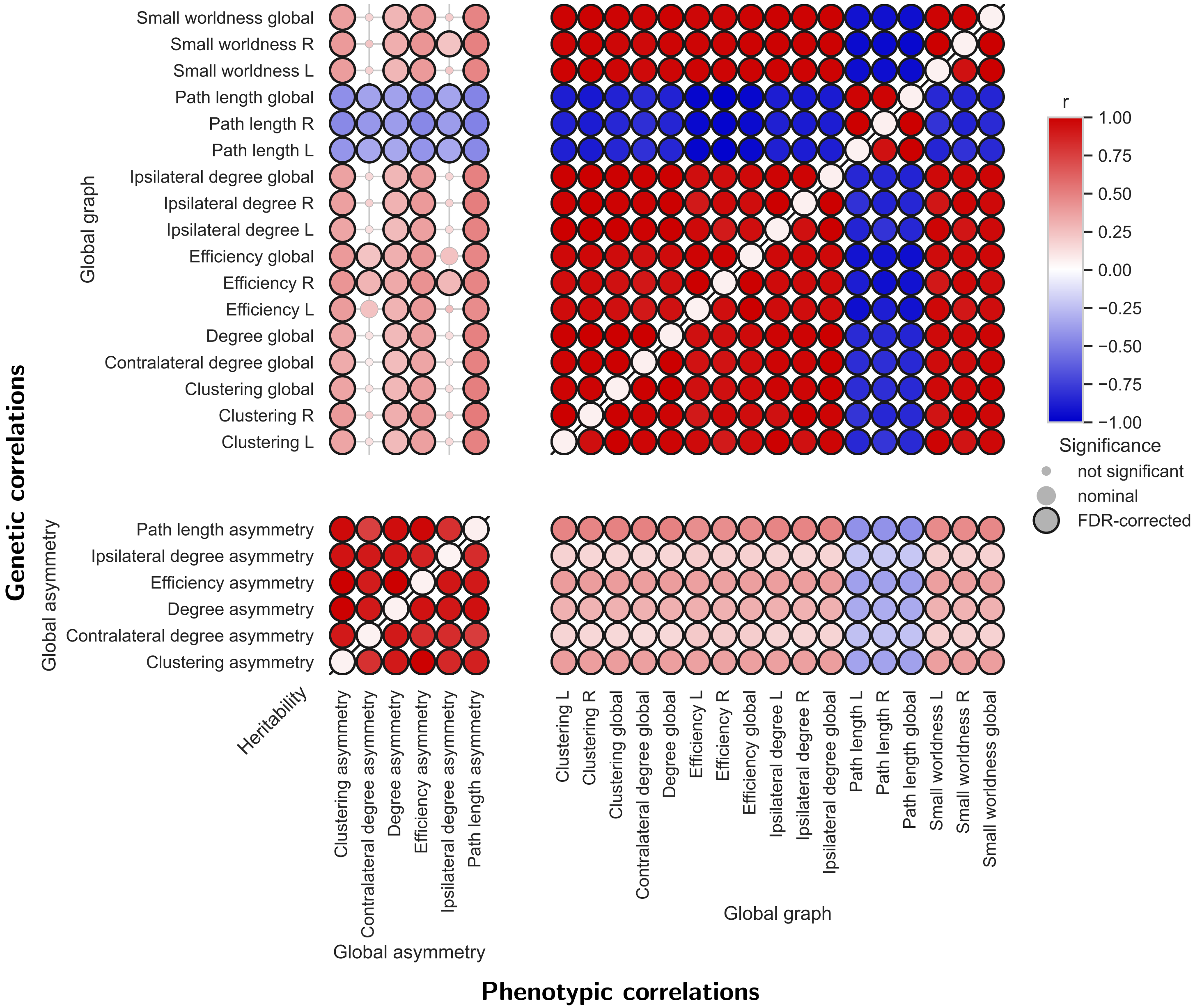


Supplementary Figure 4: **Heritability, phenotypic and genetic correlations across ten global graph phenotypes and asymmetry phenotypes.** The upper triangle depicts genetic correlations, the lower triangle depicts phenotypic correlations and the marked diagonal indicates the heritability estimated by LDSC. FDR-significant correlations/heritability are indicated with dark borders. Overall, two closely correlated clusters of traits are observed, one across asymmetry phenotypes and one across glboal and hemispheric network phenotypes.

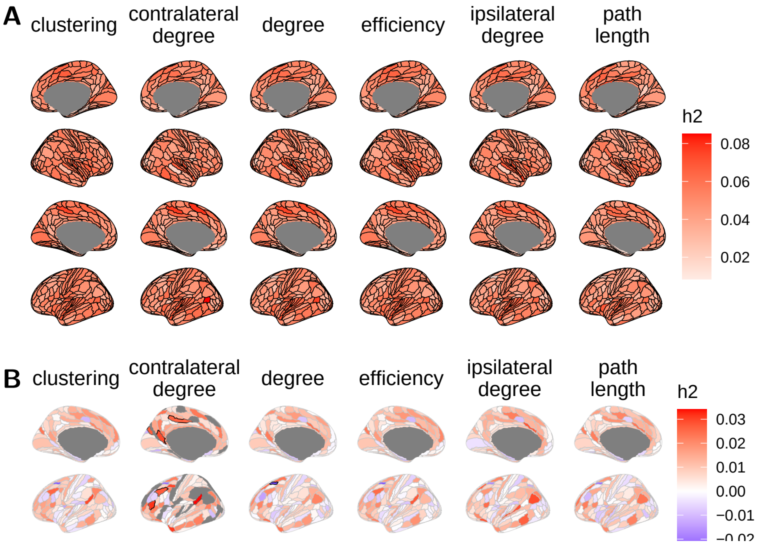


Supplementary Figure 5: **Heritability of regional phenotypes.** A. Heritability of regional graph phenotypes as estimated by LDSC. FDR-significant regions are highlited by dark borders. Most regions have significant heritability. B. LDSC-estimated Heritability of regional asymmetry using the fractional measure. Because only a few regions are significantly heritable, we did not further analyse regional-level asymmetry.

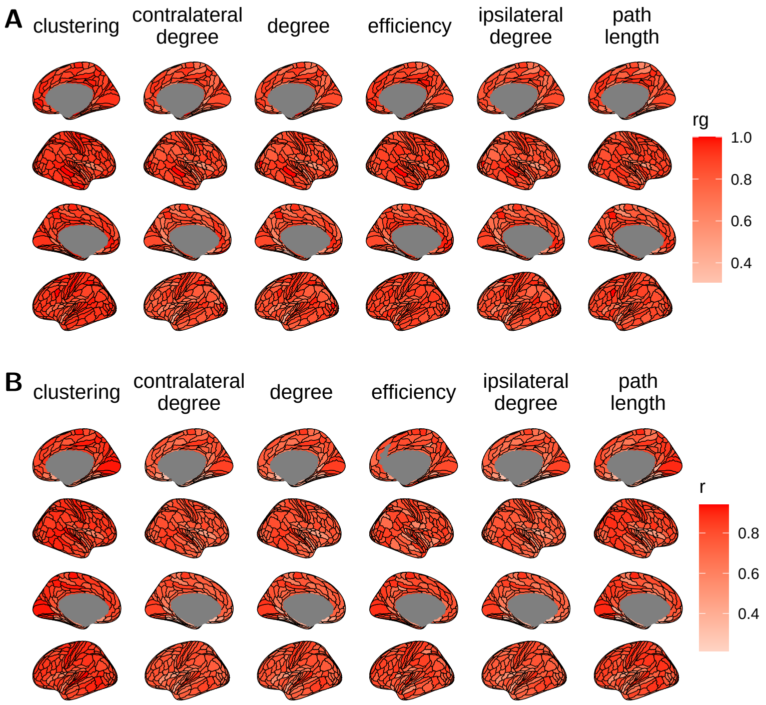


Supplementary Figure 6: **Correlation between regional graph phenotypes and corresponding global graph phenotype.** A. Genetic correlation. B. Phenotypic correlation. FDR-significant correlations are highlighted with dark borders.

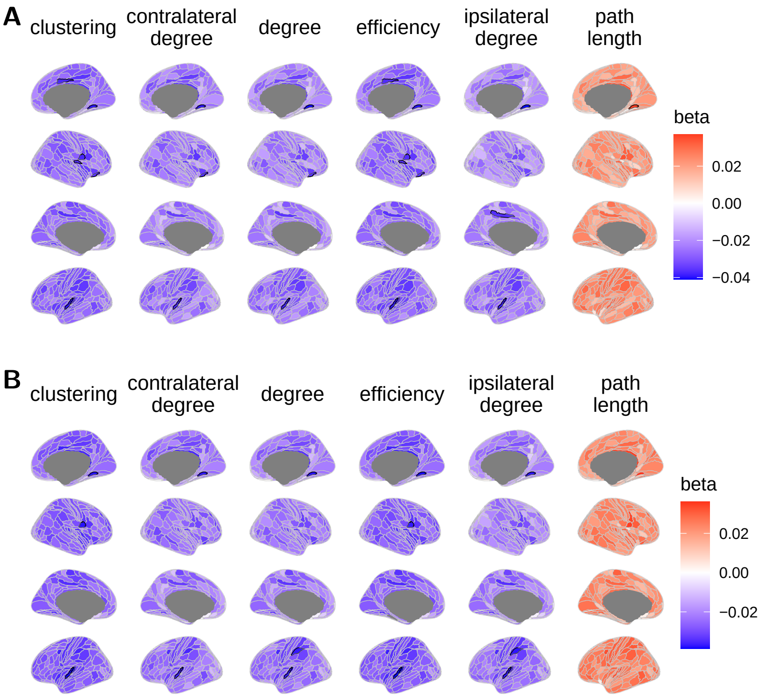


Supplementary Figure 7: **SNP-level effect sizes on regional phenotypes.** A. Effect sizes for rs2735103. B. Effect sizes for rs9393989. Genome-wide associations ($p <$ 5e-08) are highlighted with dark borders.

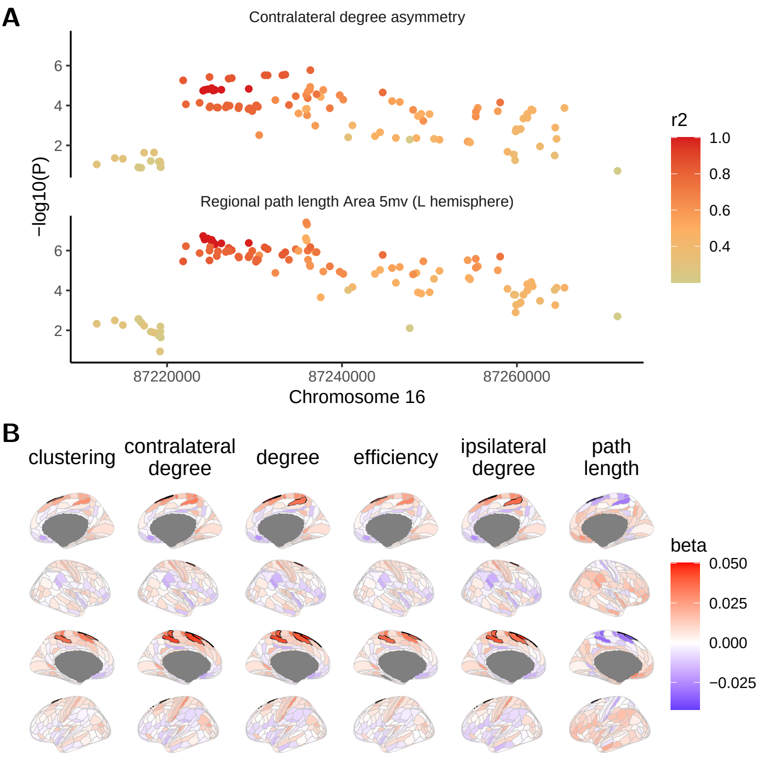


Supplementary Figure 8: **SNP-level effect sizes for rs12711472.** A. Locuszoom plot for rs12711472. Each point represents one SNP, the X axis is the genomic position, the Y axis is the significance level, and the colour indicates the linkage disequilibrium between the SNP and rs12711472. B. Effect sizes for rs12711472 on regional graph phenotypes. Genome-wide associations ($p <$ 5e-08) are highlighted with dark borders.

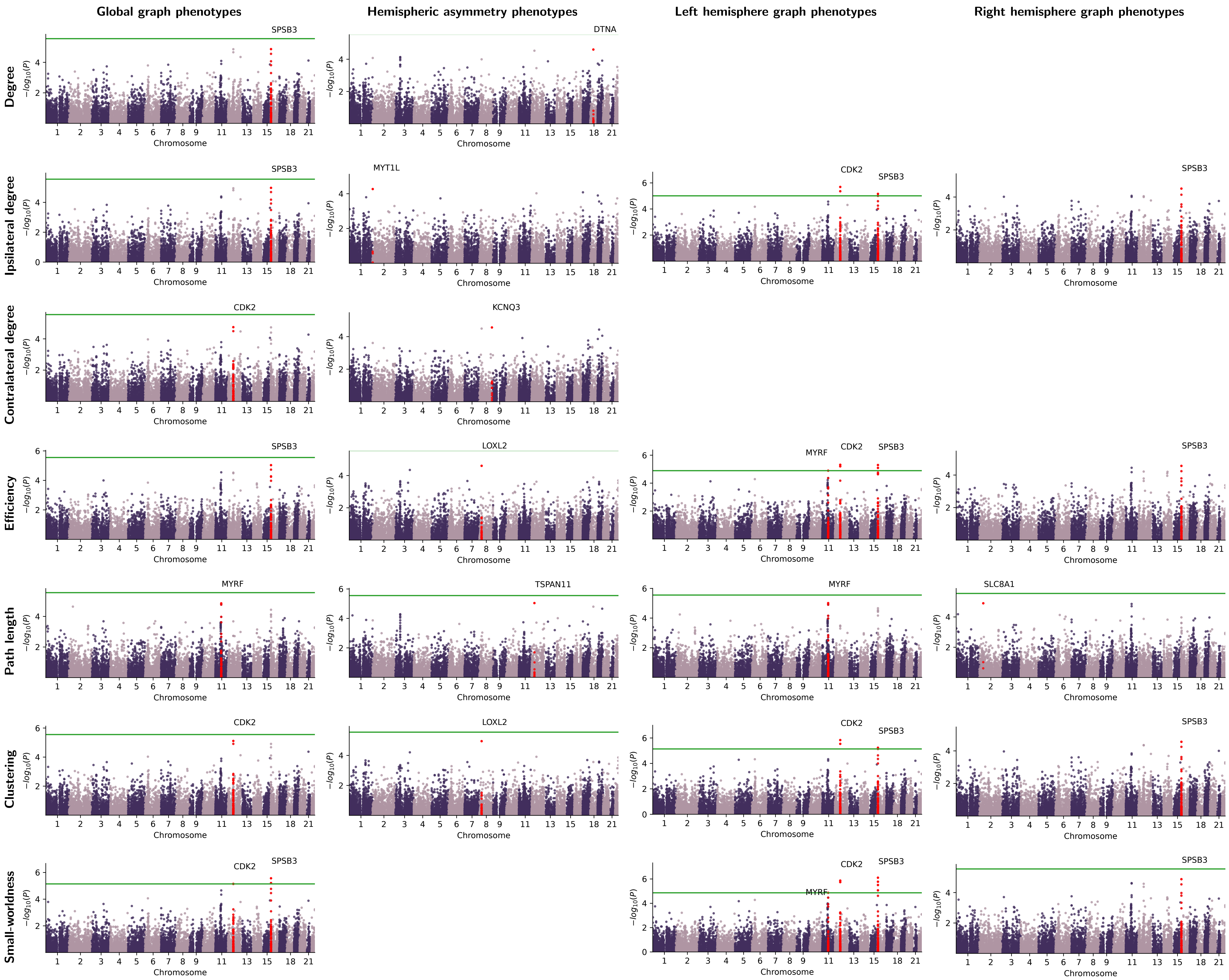


Supplementary Figure 9: **Gene-level Manhattan plot for all global, hemispheric and asymmetry phenotypes using MAGMA.** Effect sizes and p-values were obtained by aggregating SNP-level summary statistics from 35kb upstream to 10kb downstream of a gene with MAGMA. Each point represents a gene, and the y axis indicates its p-value in log scale. The green line represents the genome-wide $p_{fdr} = 0.05$ signifcance threshold. Genes within ±1Mb from the top gene are coloured red.

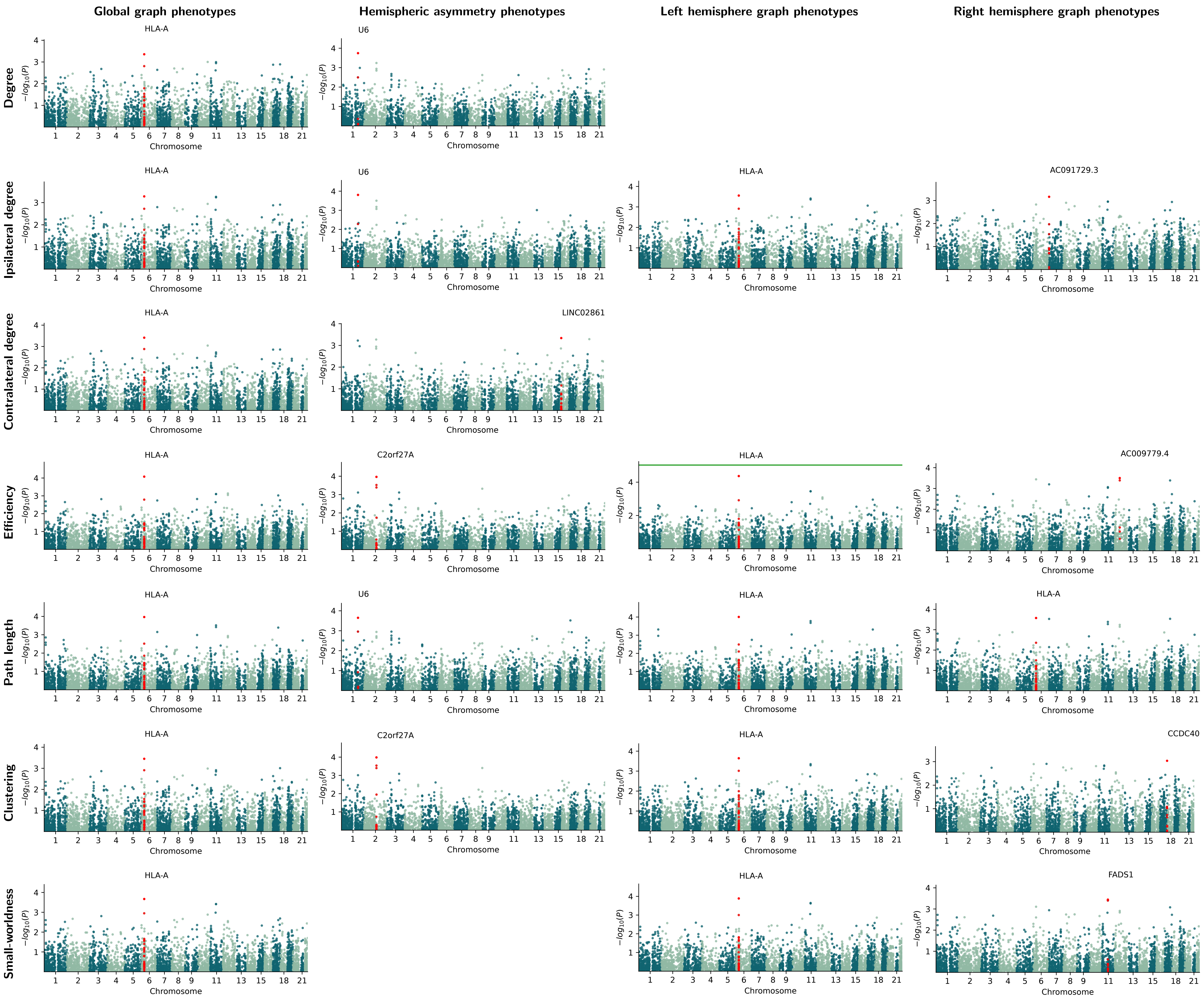


Supplementary Figure 14: **Gene-level Manhattan plot for all global, hemispheric and asymmetry phenotypes using expression quantitative trait loci (eQTL) data from the foetal brain.** Effect sizes of each gene onto phenotypes were estimated using summary-data Mendelian randomisation (SMR). Each point represents a gene, and the y axis indicates its p-value in log scale. The green line represents the genome-wide $p_{fdr} = 0.05$ signifcance threshold. Genes within ±1Mb from the top gene are coloured red.

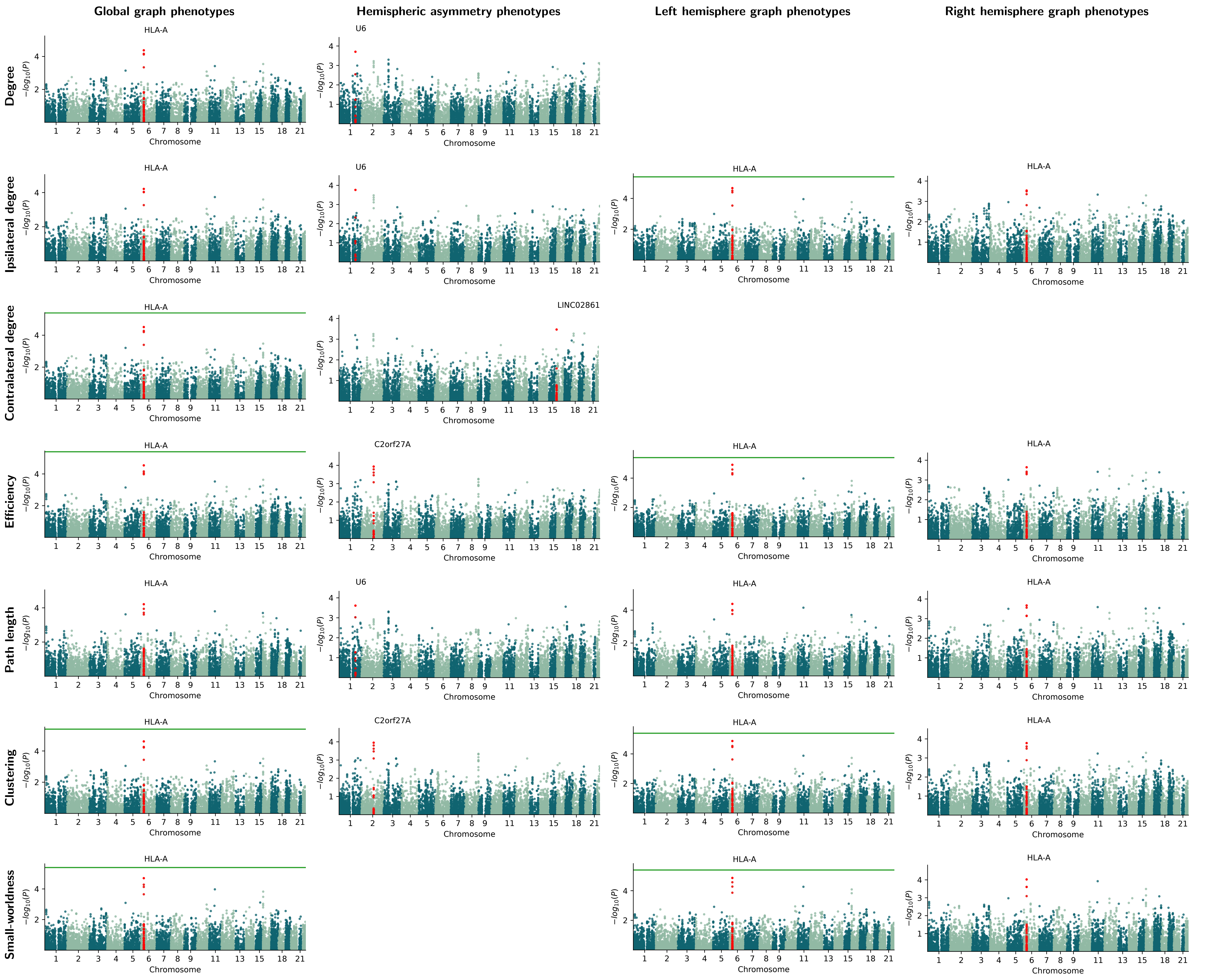


Supplementary Figure 15: **Gene-level Manhattan plot for all global, hemispheric and asymmetry phenotypes using isoform quantitative trait loci (isoQTL) data from the foetal brain.** Effect sizes of each gene onto phenotypes were estimated using summary-data Mendelian randomisation (SMR). Each point represents a gene, and the y axis indicates its p-value in log scale. The green line represents the genome-wide $p_{fdr} = 0.05$ signifcance threshold. Genes within ±1Mb from the top gene are coloured red.

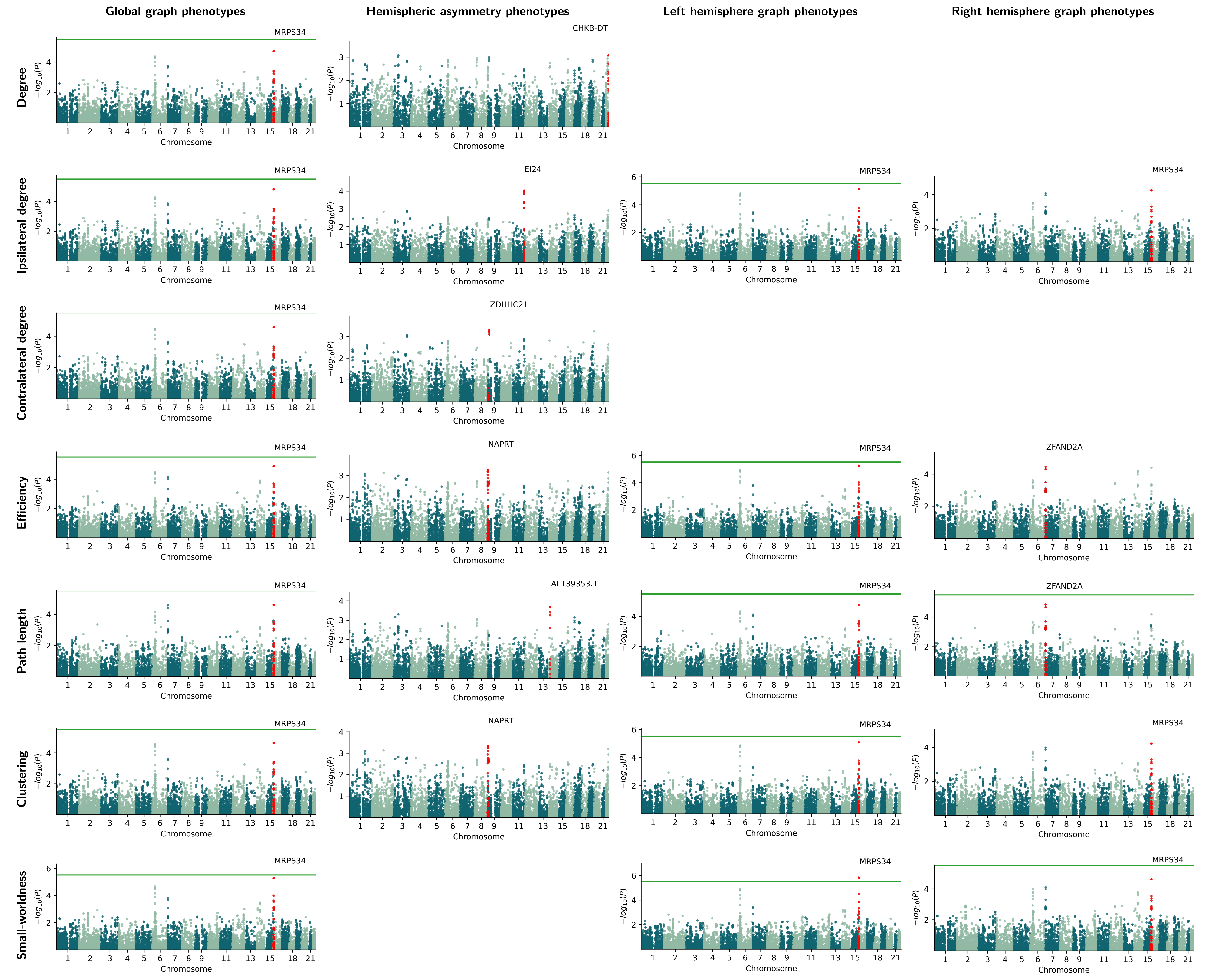


Supplementary Figure 16: **Gene-level Manhattan plot for all global, hemispheric and asymmetry phenotypes using splicing quantitative trait loci (sQTL) data from the foetal brain.** Effect sizes of each gene onto phenotypes were estimated using summary-data Mendelian randomisation (SMR). Each point represents a gene, and the y axis indicates its p-value in log scale. The green line represents the genome-wide $p_{fdr} = 0.05$ signifcance threshold. Genes within ±1Mb from the top gene are coloured red.

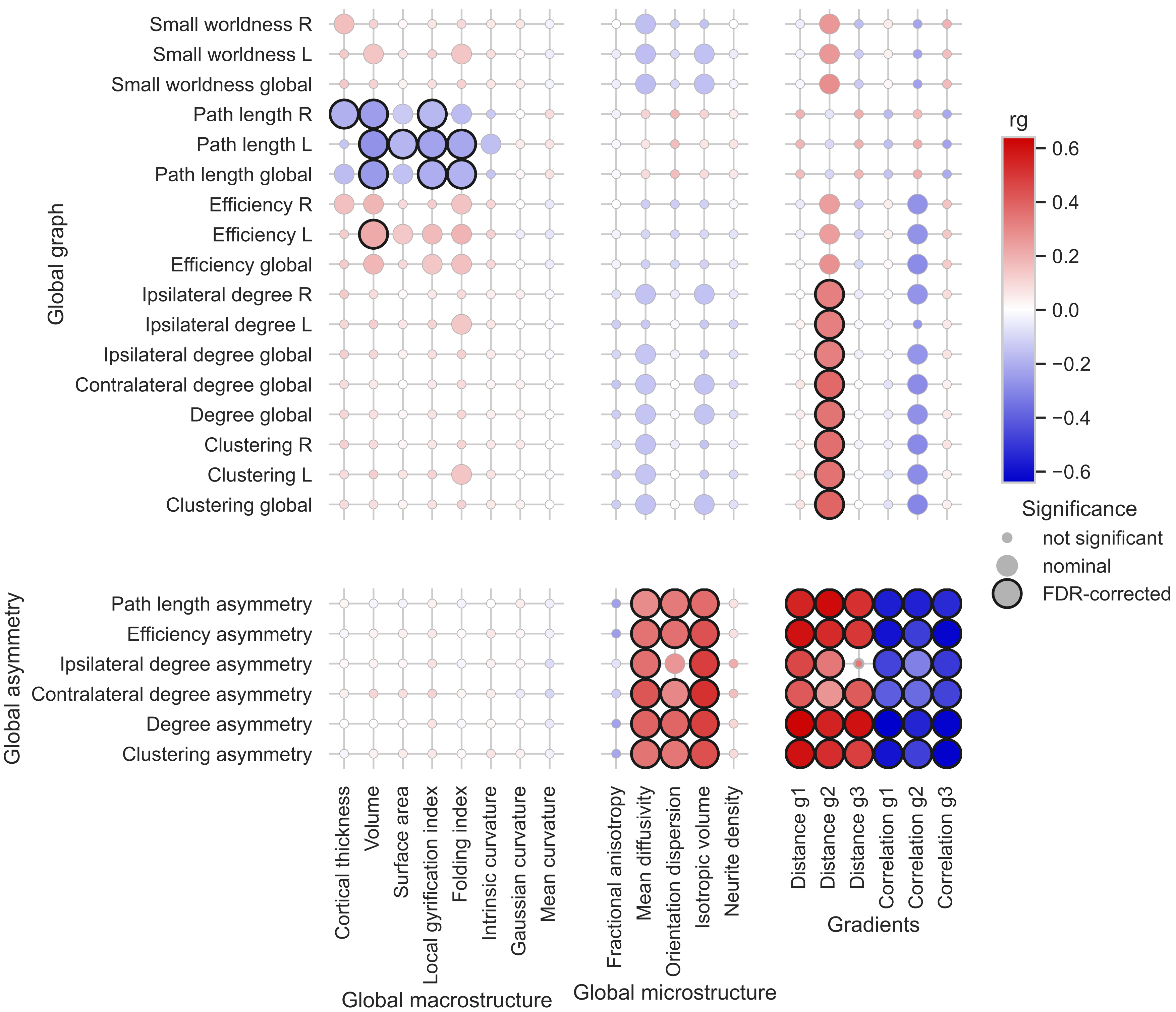


Supplementary Figure 17: **Genetic correlation between graph phenotypes and other imaging derived phenotypes.** Colour scale represents LDSC-estimated genetic correlation. Significant correlations after FDR correction are highlighted with dark borders. G1-G3 are the top three functional gradients, and the similarity between the regional gradiant values with the population template of HCP were used to quantify the individual difference in the functional gradient hierarchy. Two different parametrisations were used, i.e. Euclidean distance and Pearson's correlation.

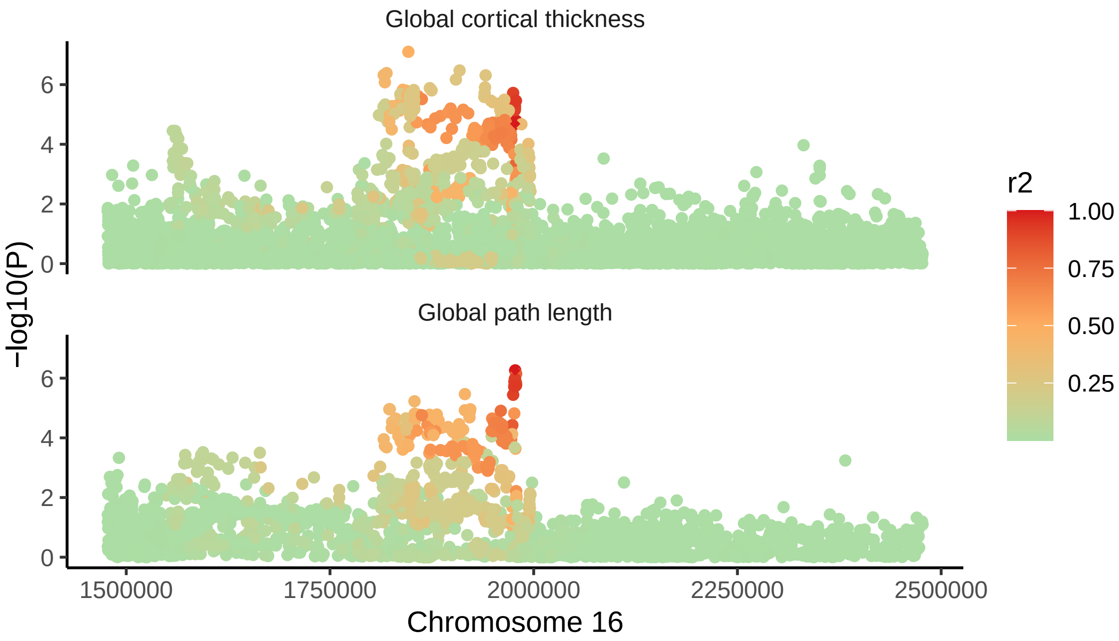


Supplementary Figure 18: **Summary statistics for the region rs186940 ± 1 Mb (chromosome 16, 0.9-2.9 Mb)** showed colocalisation between global graph phenotypes and cortical thickness. Each point represents a SNP, the y axis indicates the GWAS p-value in log scale and the colour scale represents its linkage disequilibrium with the candidate SNP (rs186940).

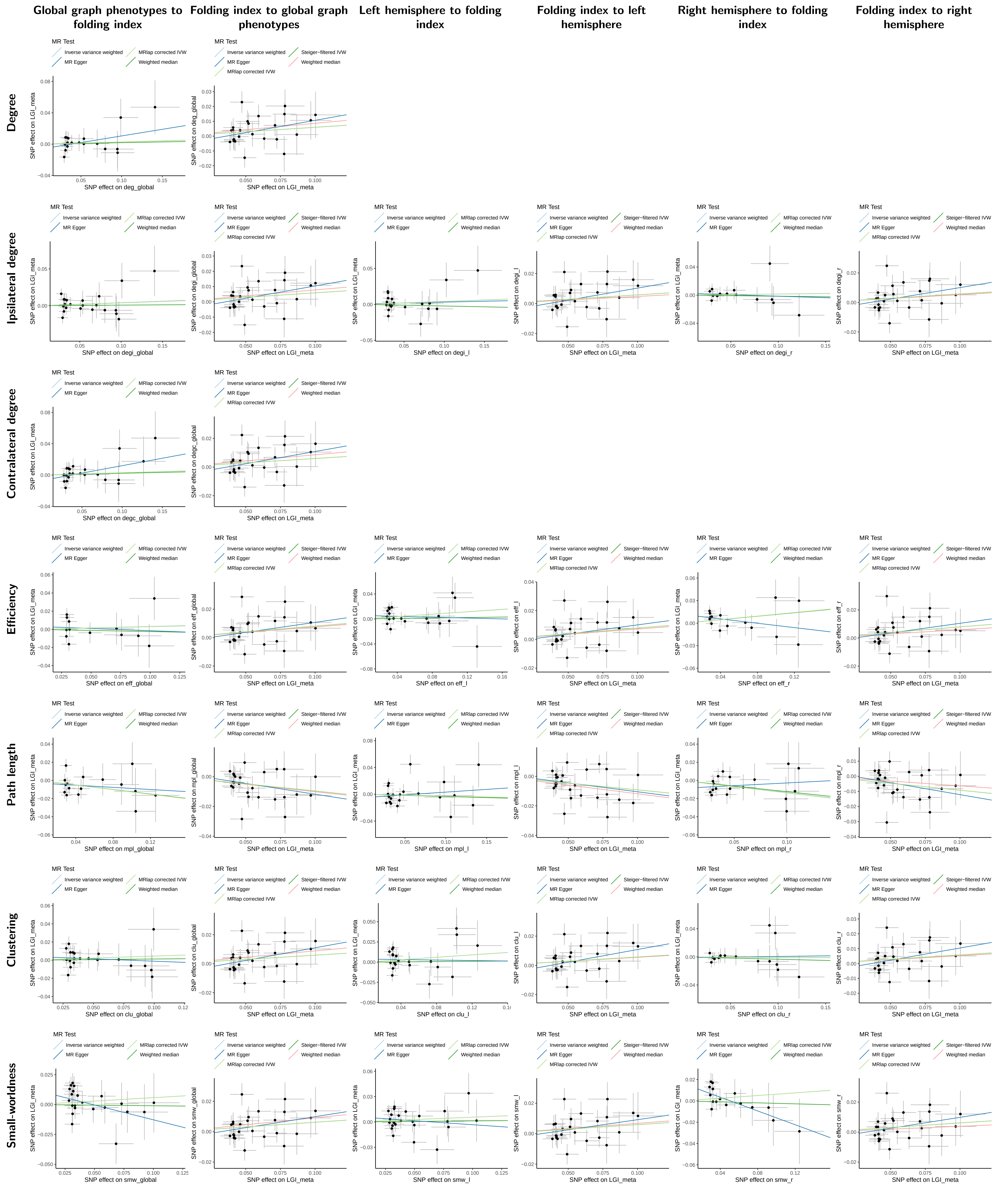


Supplementary Figure 19: **Mendelian randomisation scatter plot examining the causality between global graph phenotypes and local gyrification index**, with estimated fits using five different methods. Each dot represents a SNP, and its x and y axis positions indicate its GWAS-estimated effect size on global path length and cortical volume. Error bars for each dot indicate its GWAS-estimated standard error. Only significant SNPs were included (see Methods).

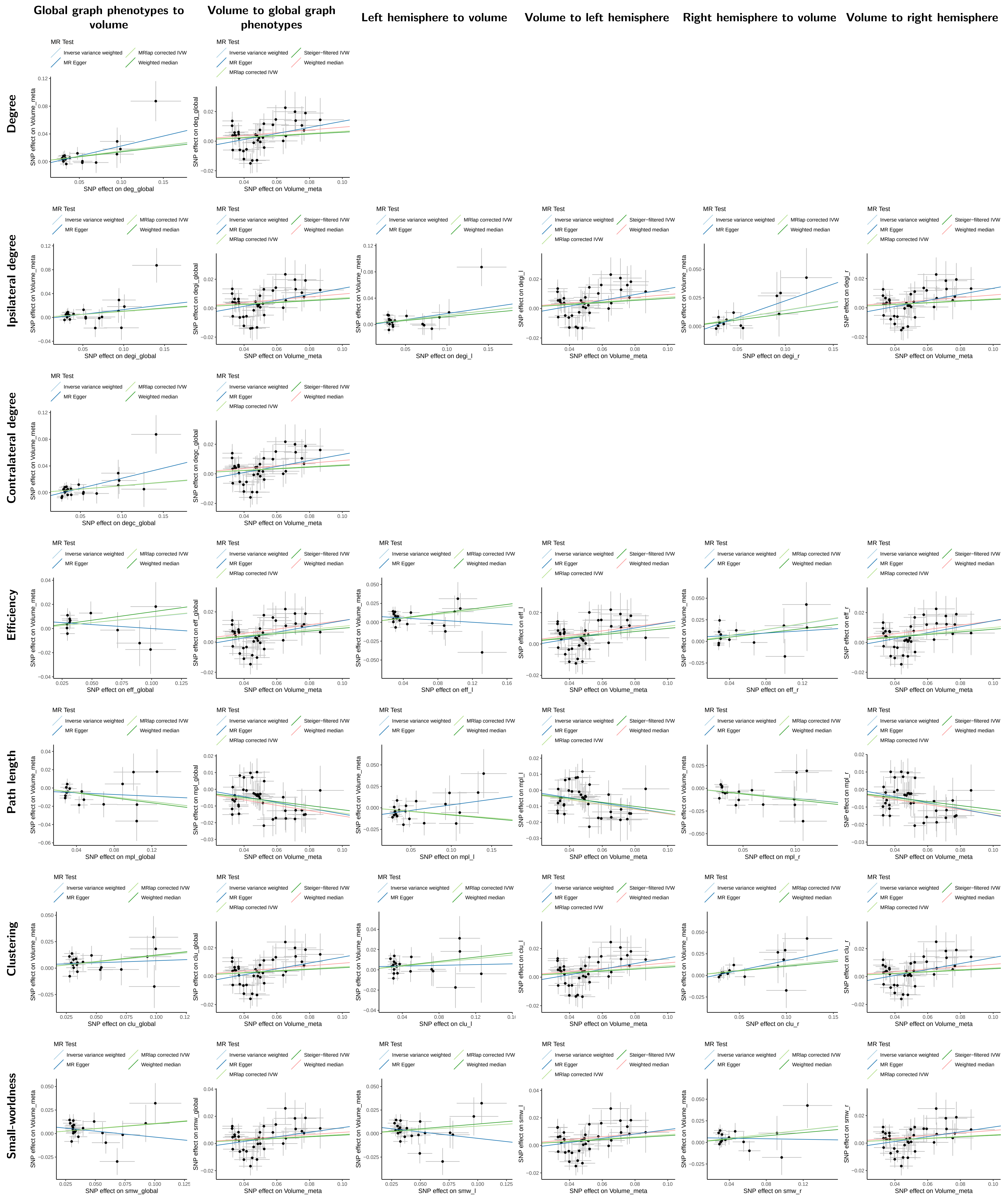


Supplementary Figure 20: **Mendelian randomisation scatter plot examining the causality between global graph phenotypes and cortical volume,** with estimated fits using five different methods. Each dot represents a SNP, and its x and y axis positions indicate its GWAS-estimated effect size on global path length and cortical volume. Error bars for each dot indicate its GWAS-estimated standard error. Only significant SNPs were included (see Methods).

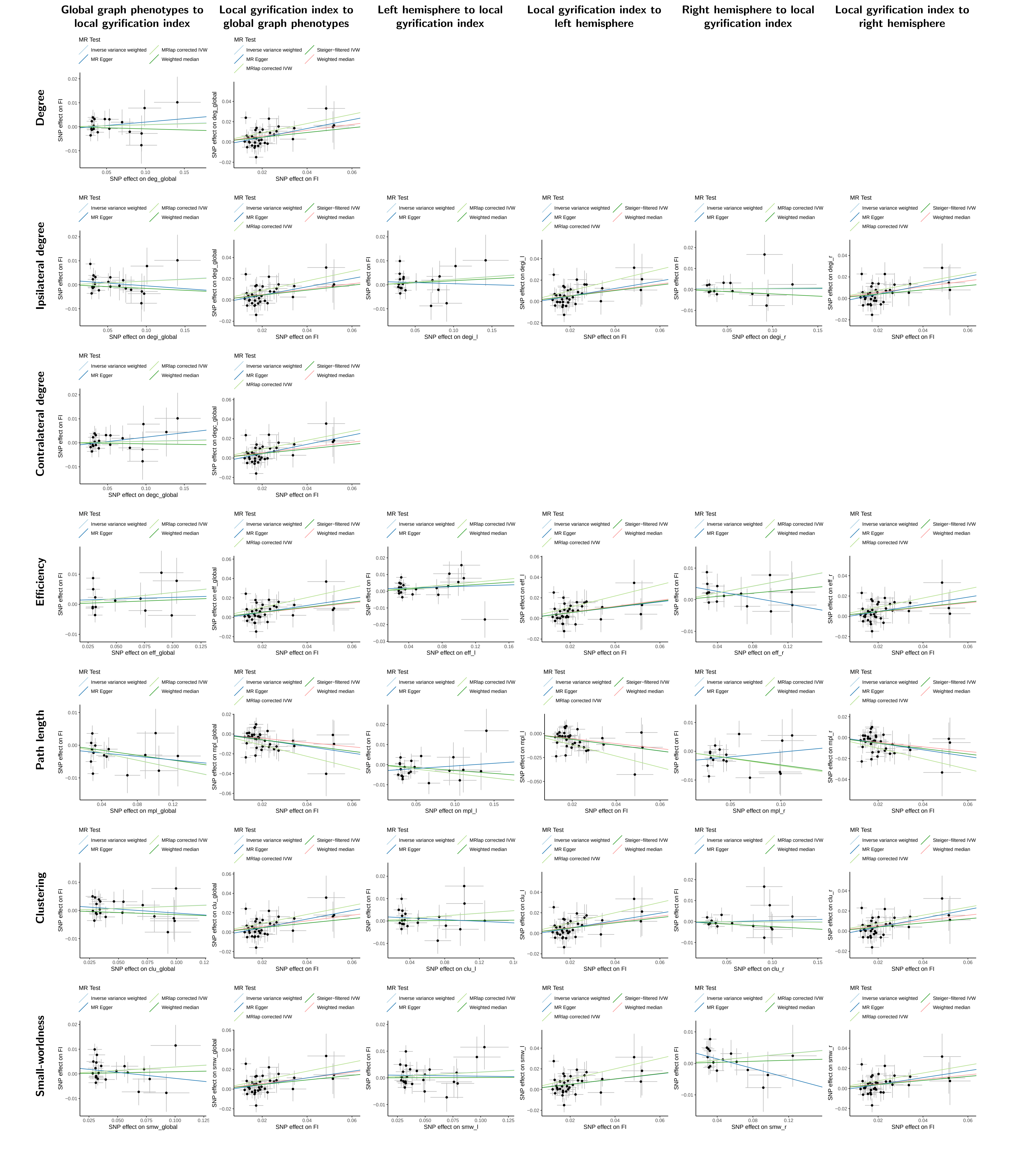


Supplementary Figure 21: **Mendelian randomisation scatter plot examining the causality between global graph phenotypes and cortical folding index**, with estimated fits using five different methods. Each dot represents a SNP, and its x and y axis positions indicate its GWAS-estimated effect size on global path length and cortical volume. Error bars for each dot indicate its GWAS-estimated standard error. Only significant SNPs were included (see Methods).

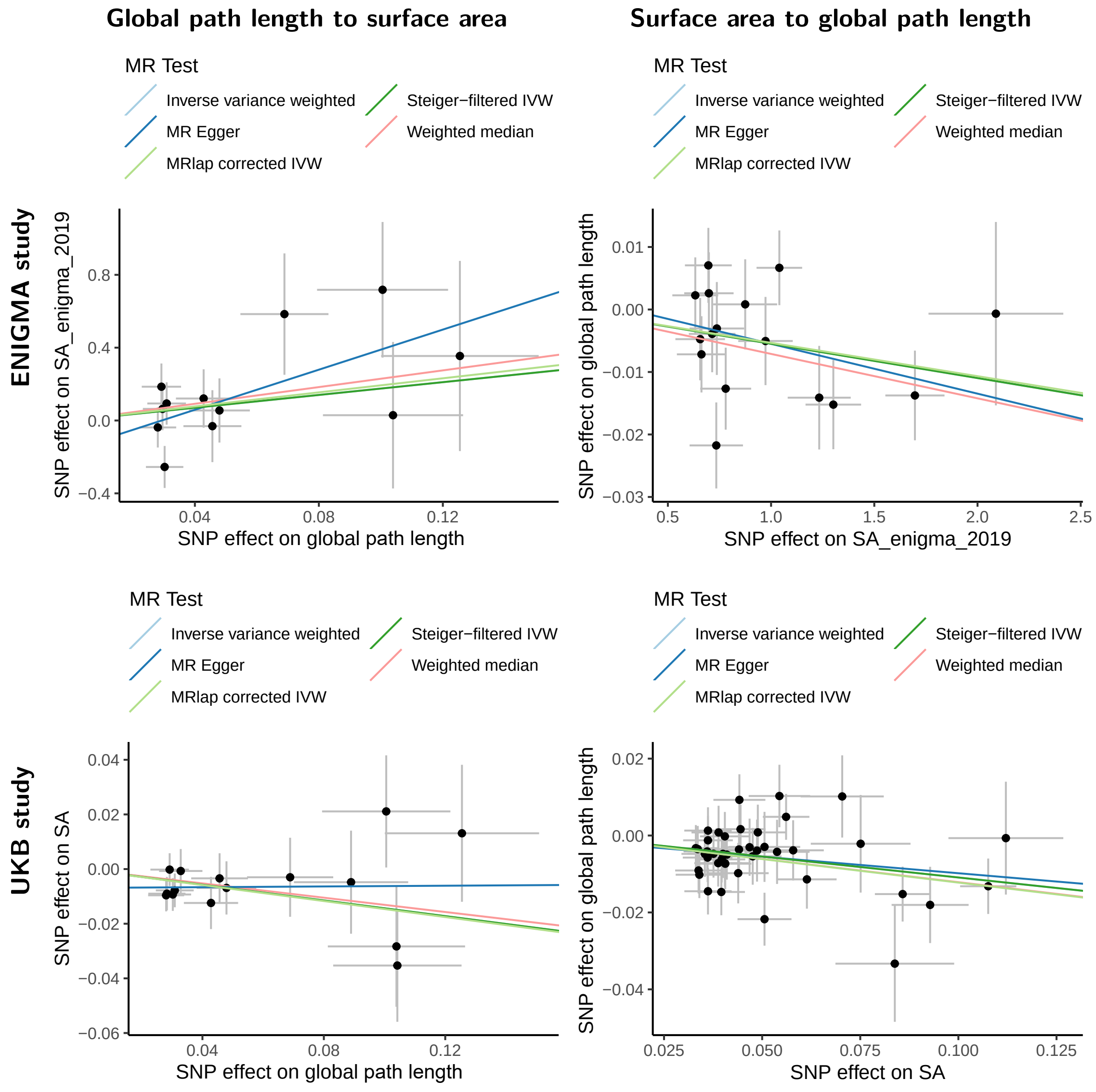


Supplementary Figure 22: **Mendelian randomisation scatter plot examining the causality between global path length and surface area in the ENIGMA study, compared with the UKB 2023 study**, with estimated fits using five different methods. Each dot represents a SNP, and its x and y axis positions indicate its GWAS-estimated effect size on global path length and cortical volume. Error bars for each dot indicate its GWAS-estimated standard error. Only significant SNPs were included (see Methods).

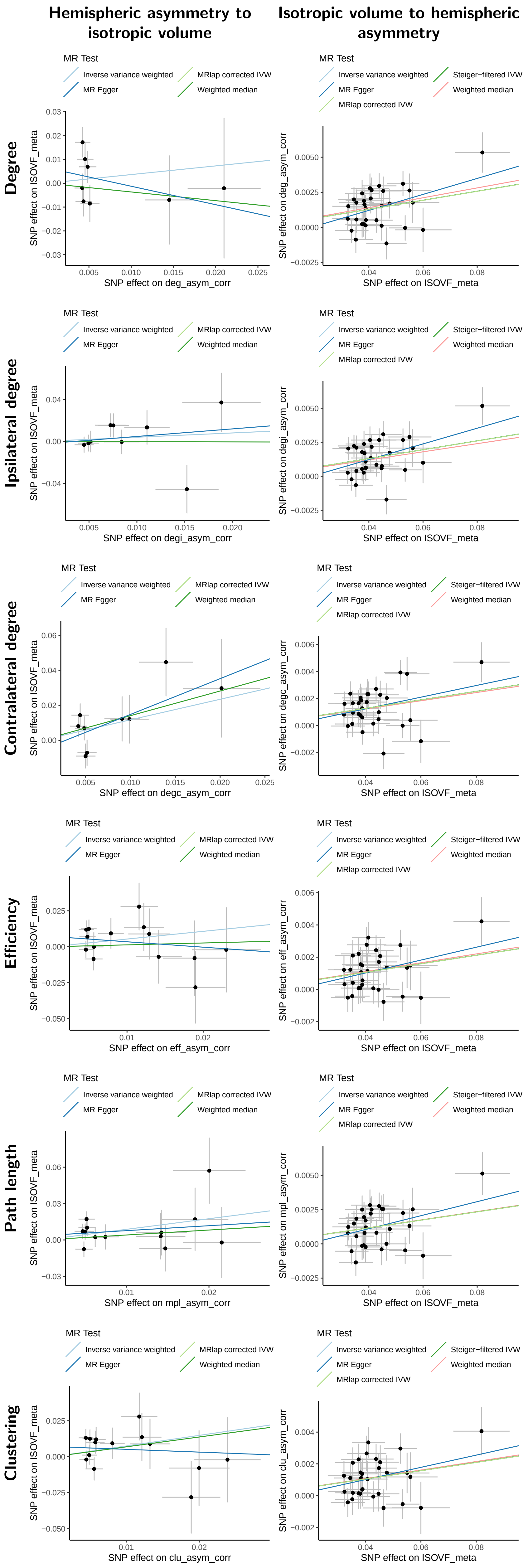


Supplementary Figure 23: **Mendelian randomisation scatter plot examining the causality between asymmetry phenotypes and isotropic volume,** with estimated fits using five different methods. Each dot represents a SNP, and its x and y axis positions indicate its GWAS-estimated effect size on global path length and cortical volume. Error bars for each dot indicate its GWAS-estimated standard error. Only significant SNPs were included (see Methods).

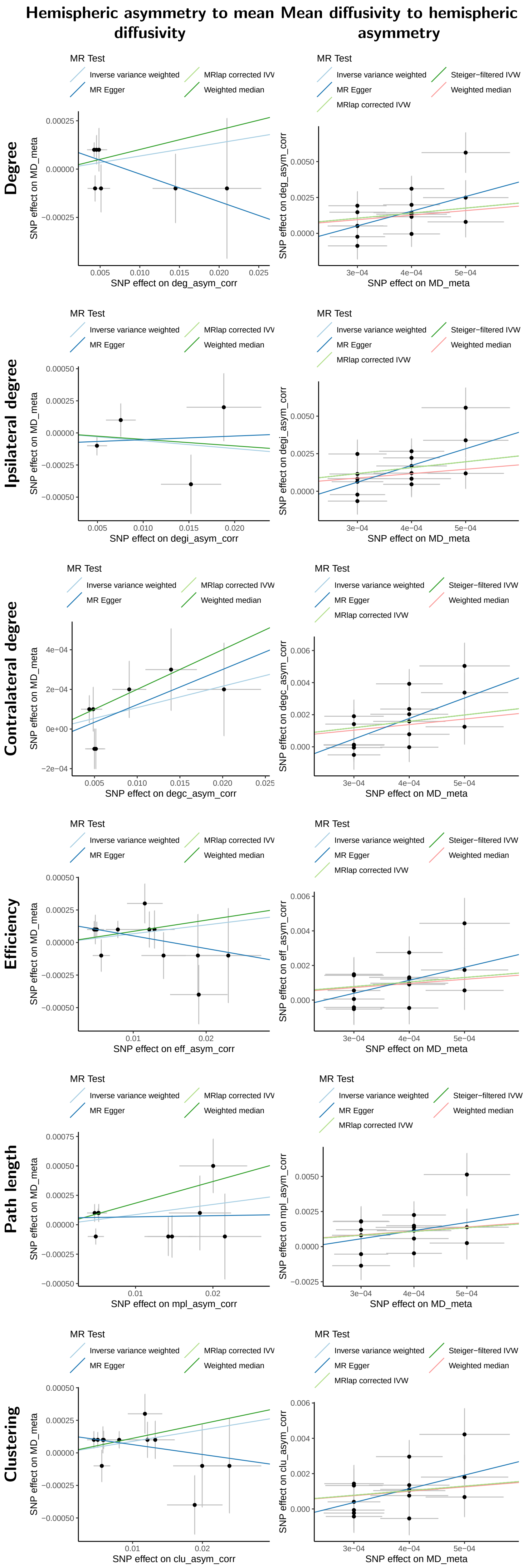


Supplementary Figure 24: **Mendelian randomisation scatter plot examining the causality between asymmetry phenotypes and mean diffusivity,** with estimated fits using five different methods. Each dot represents a SNP, and its x and y axis positions indicate its GWAS-estimated effect size on global path length and cortical volume. Error bars for each dot indicate its GWAS-estimated standard error. Only significant SNPs were included (see Methods).

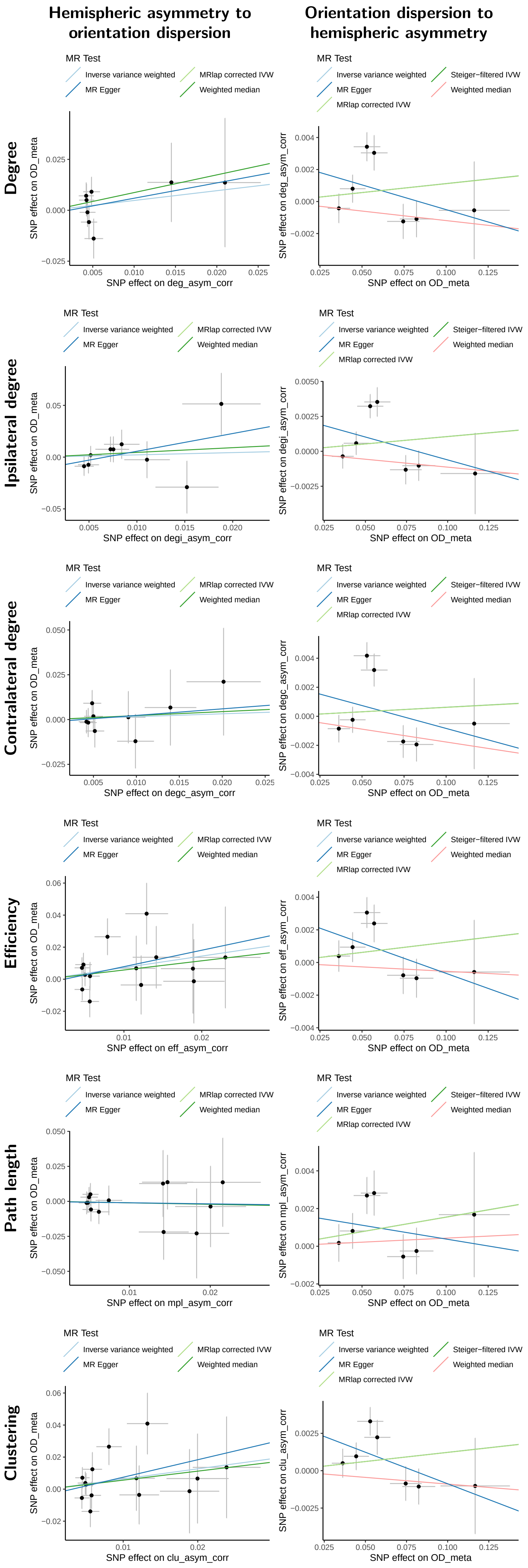


Supplementary Figure 25: **Mendelian randomisation scatter plot examining the causality between asymmetry phenotypes and orientation dispersion index**, with estimated fits using five different methods. Each dot represents a SNP, and its x and y axis positions indicate its GWAS-estimated effect size on global path length and cortical volume. Error bars for each dot indicate its GWAS-estimated standard error. Only significant SNPs were included (see Methods).

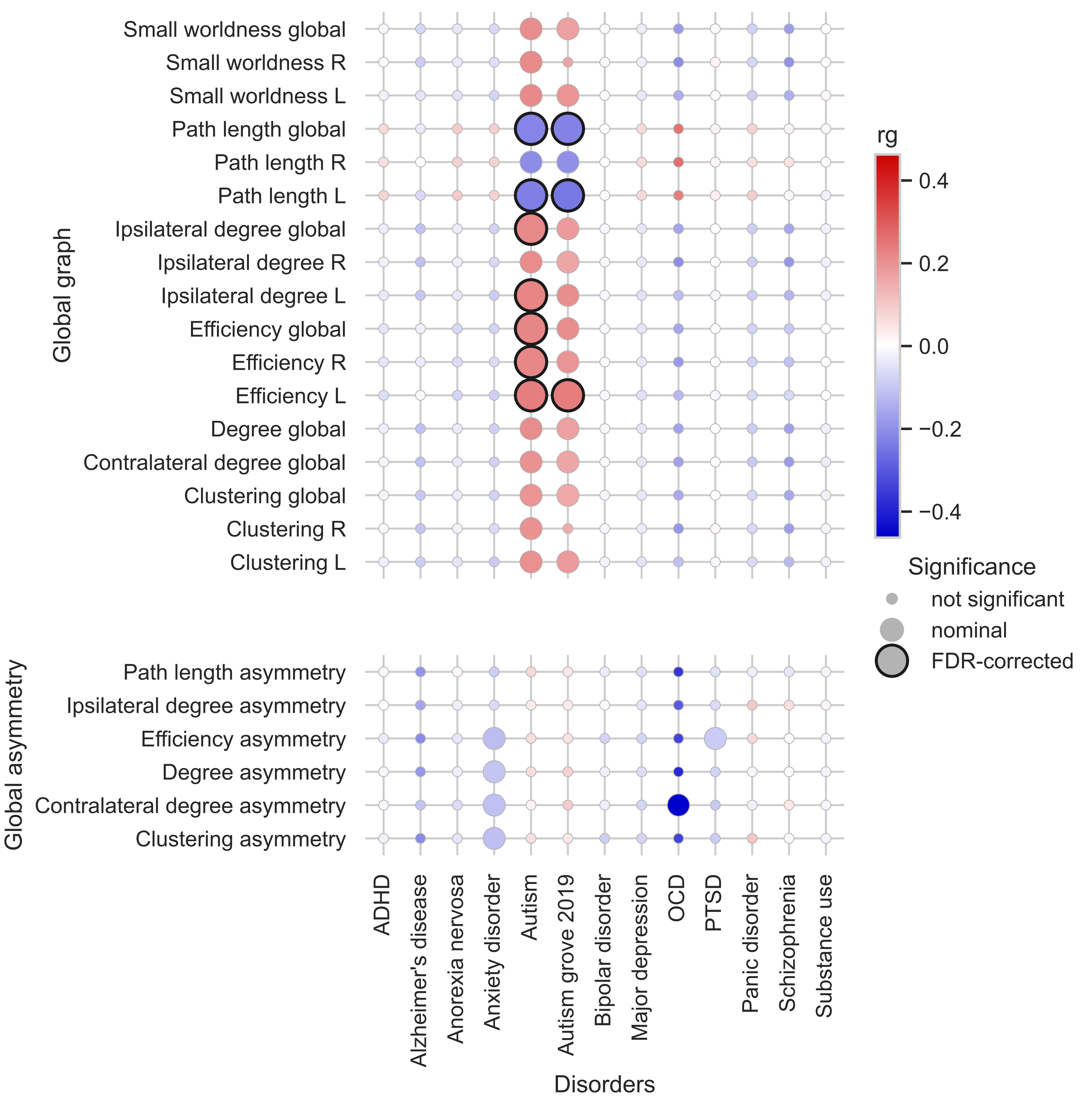


Supplementary Figure 26: **Genetic correlation between global and hemispheric graph phenotypes, hemispheric asymmetry and 12 neuropsychiatric disorders**, estimated with LDSC. Colour scale represents the genetic correlation coefficient and significant correlations after FDR correction are highlighted with dark borders. ADHD: attention-deficit hyperactivity disorder. MDD: major depressive disorder. OCD: obsessive-compulsive disorder. PTSD: post-trauma stress disorder. SUD: substance use disorder.

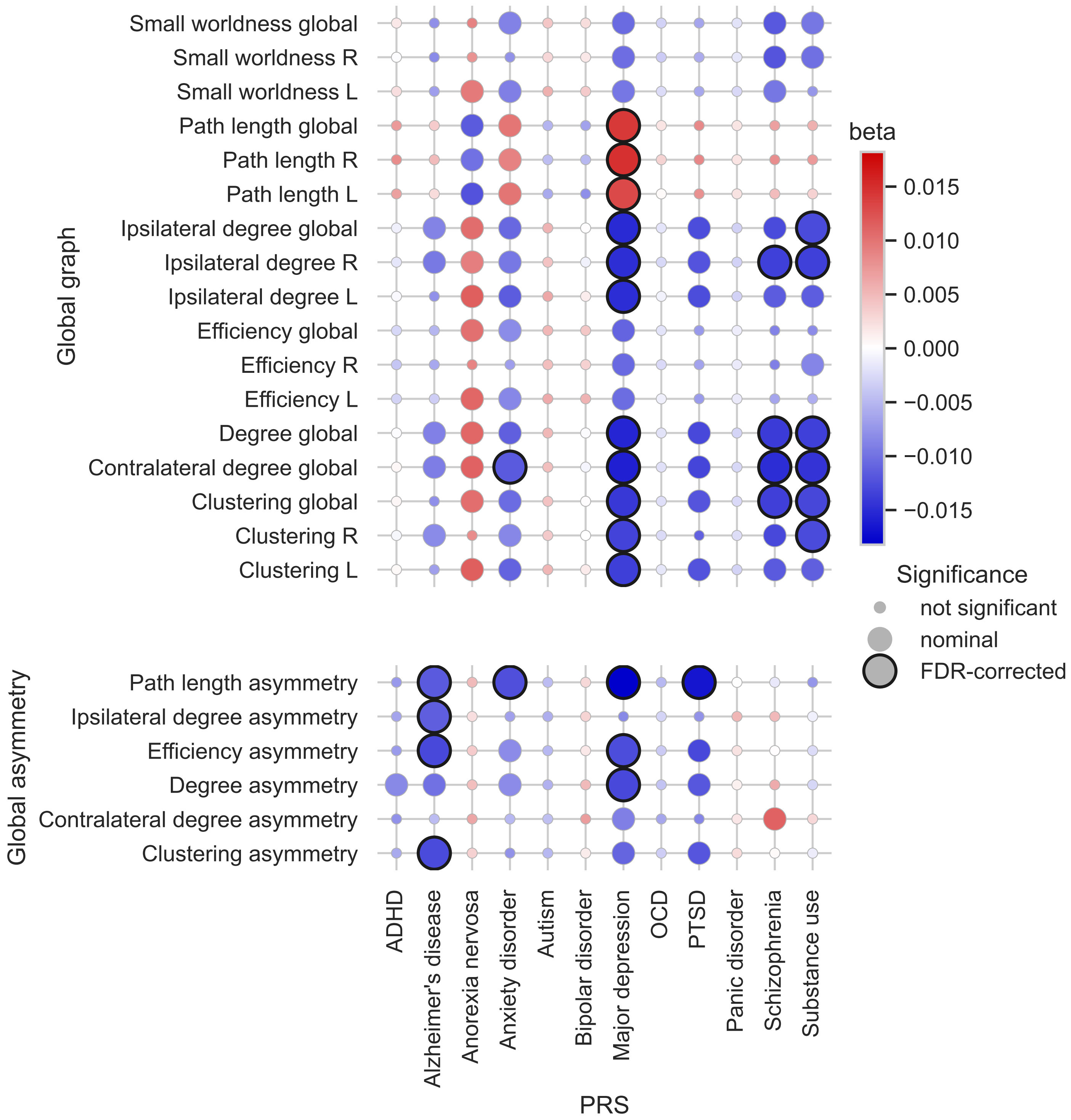


Supplementary Figure 27: **Polygenic score based associations between graph phenotypes and neuropsychiatric disorders.** Polygenic scores for disorders were estimated using PRScs. Then, the effect of disorder polygenic score on graph phenotypes was estimated using linear modelling, with the same covariates as the GWAS analysis (see Methods). Colour scale represents the effect size and significant associations after FDR correction are highlighted with dark borders. ADHD: attention-deficit hyperactivity disorder. MDD: major depressive disorder. OCD: obsessive-compulsive disorder. PTSD: post-trauma stress disorder. SUD: substance use disorder.

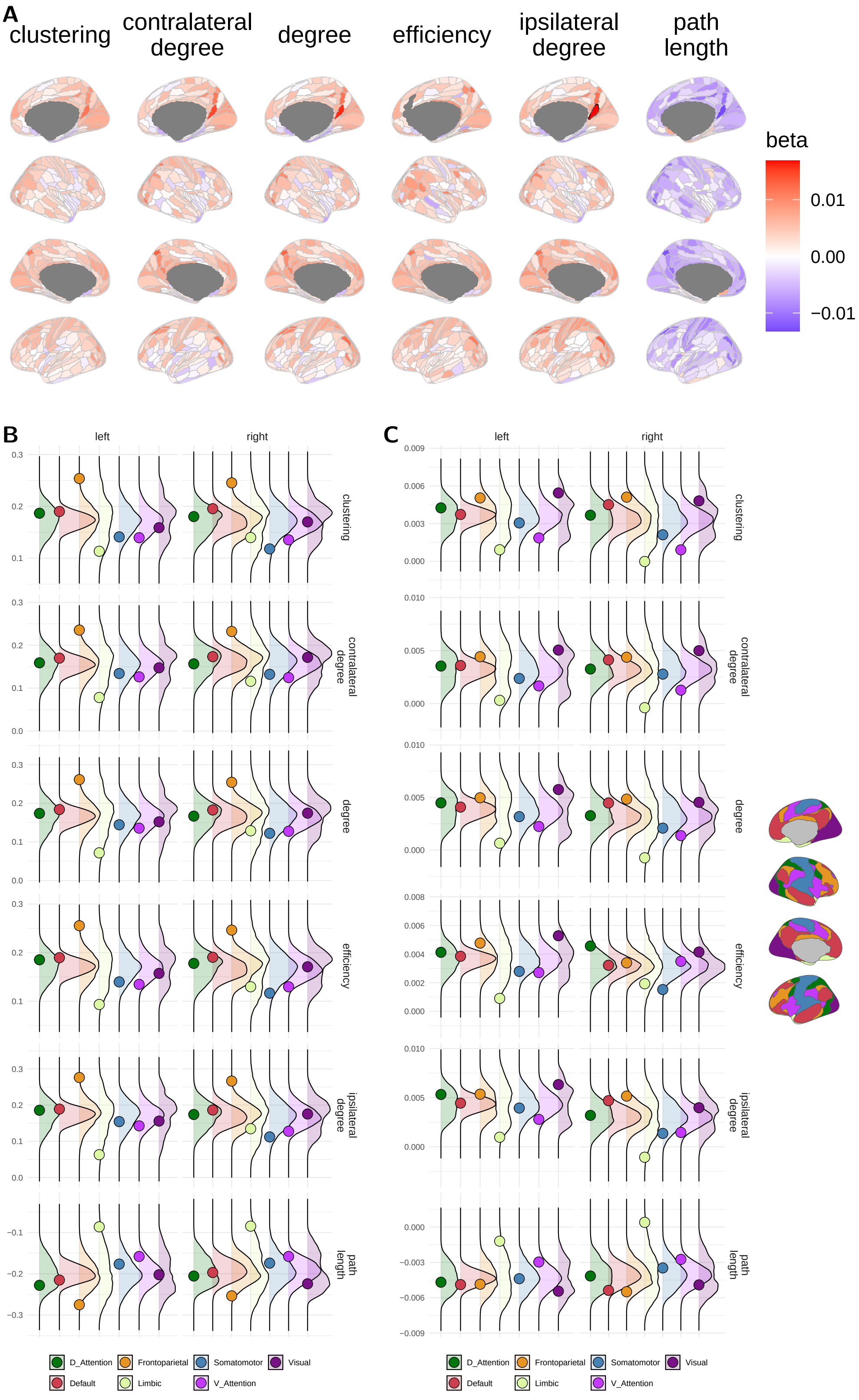


Supplementary Figure 28: **Association between regional graph phenotypes with polygenic score of autism.** A. Linear-modelling based effect size of polygenic score on regional graph phenotypes. Significant associations are highlighted with dark borders. B-C. Spherical spin-permutation test results for genetic correlation (B) and polygenic score-based association (C) between regional graph phenotypes and autism. For each functional network of the Yeo-7 atlas, the coloured dot represents the mean genetic correlation with autism across all regions within that network, and the curve represents the spherically permuted null-distribution. The graphical legend indicates the spatial positioning of the Yeo-7 networks.

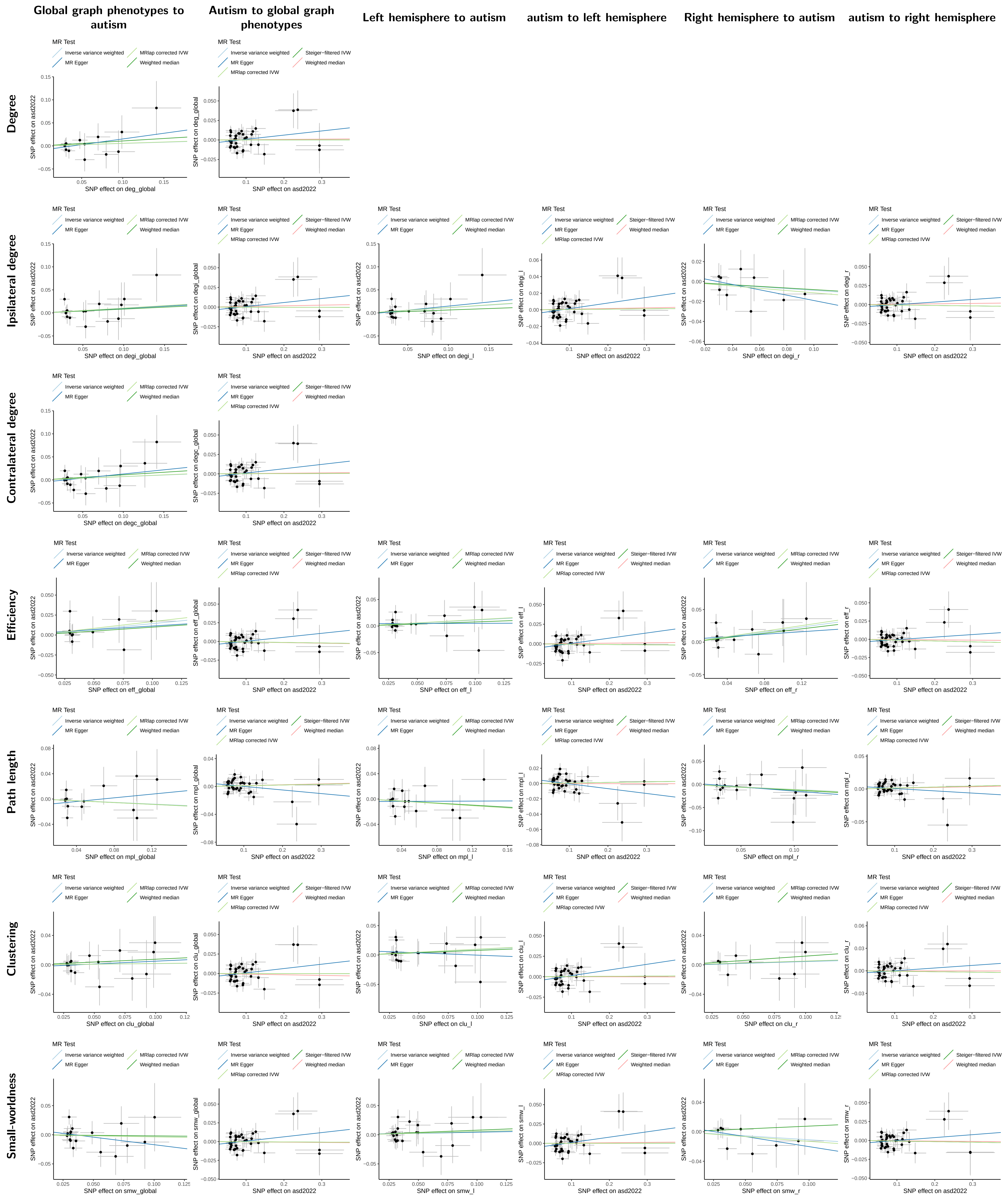


Supplementary Figure 29: **Mendelian randomisation scatter plot examining the causality between global graph phenotypes and autism**, with estimated fits using five different methods. Each dot represents a SNP, and its x and y axis positions indicate its GWAS-estimated effect size on global path length and cortical volume. Error bars for each dot indicate its GWAS-estimated standard error. Only significant SNPs were included (see Methods).

### SN1 Comparison between our results with previous functional network GWAS

Two studies investigated the common variant genetics of graph phenotypes derived from the resting-state functional network [1, 2]. Whilst both studies analysed graph phenotypes of the functional network, we note differences in functional network parametrisation before deriving the graph metrics, which may affect the genetic results. The detailed differences are outlined below.

The Foo et al. 2021 [1] study used full Pearson correlations and Fisher transform to generate the functional connectome. Negative connections and self-connections were set to zero, same as our present study. The only differences in the phenotyping protocol were: (1) Foo et al. used a coarser parcellation atlas than the present study; (2) we normalised the connectivity strengths to [0,1] as required to measure clustering [3, 4], whereas the Foo et al. 2021 study did not. As a sensitivity analysis, we derived the same graph phenotypes (except for clustering) using the raw connectivity strengths without normalisation. Phenotypic correlations between the two approaches were perfect for asymmetry (because Spearman correlation was used for asymmetry phenotypes), very high for degree phenotypes and efficiency (minimum r = 0.863 across all regions), and also fairly high for path length (global r = 0.644, minimum r = 0.639 and mean r = 0.796 across all regions, Supplementary Table 2.13). Because we replicated the significant locus identified by Foo et al. 2021 at a high significance level (p = 6e-07 to 7e-04, Supplementary Table 1.2), we were confident that this difference in network parametrisation did not substantially change the results.

The Bell et al. 2022 [2] study also used full Pearson correlations and Fisher transform, but only selected the top 10% of connections to generate the connectome. All other connections were set to zero. Previous phenotypic studies have suggested that weak functional connections are also significant, for example in cognitive functioning [5]. The discrepancy between our genetic findings and the Bell et al. 2022 study (only 4 out of 7 loci in consistent direction, Supplementary Table 1.2) can possibly be explained by different genetic processes underlying weak and strong connections.

### SN2 Understanding hemispheric asymmetry of graph phenotypes

Whilst the correlative measure of hemispheric asymmetry is mathematically defined to increase with dissimilarity between hemispheric networks, genetic correlation analyses of the hemispheric asymmetry phenotypes surprisingly showed that both hemispheres were correlated in the same direction with hemispheric asymmetry phenotypes (Figure 2c). In particular, asymmetry in all phenotypes increased with total connectivity within each hemisphere. A similar pattern was identified at a regional level (Supplementary table 2.12) where corresponding regions of both hemispheres correlate in the same direction with hemispheric asymmetry. Because the population median for corresponding regional phenotypes across left and right hemispheres are generally very similar (Figure 1b), we hypothesised that hemispheric asymmetry is driven by the development of additional connectivity above a fairly symmetric baseline.

In order to test this hypothesis, we tested the phenotypic correlation between regional phenotypes and hemispheric asymmetry, adjusted for the sum of corresponding regional phenotypes in left and right hemispheres. The adjusted correlation was significantly attenuated to near zero for all six graph metrics (Supplementary table 2.13). This was consistent with the idea that the baseline connectome is fairly symmetric, and development of over-connectivity for individual regions can be more hemisphere-specific, but the direction of the hemisphere bias varies across individuals.

To provide a directional understanding on the correlation between asymmetry and global phenotypes, we conducted exploratory Mendelian Randomisation using the same methods as our main analyses (see Methods). Ample evidence suggested that global phenotypes caused hemispheric asymmetry (lowest $p_{fdr}$ = 8.94e-08) and this was supported by parallel Mendelian Randomisation methods (see Methods, $p_{fdr} < 0.05$ across three methods, Supplementary Table 3.4).

Whilst the above analysis provided initial support for the idea that over-connectivity drives functional asymmetry, it should be noted that, most clinical studies have focused on the fractional measure of asymmetry at hemisphere or regional level (which are not heritable). Hence, more investigations using the correlative definition of asymmetry are required to more thoroughly understand this phenotype.
